# Incidence of SSRI treatment and psychiatric specialist care in new-onset adult epilepsy: are newer antiseizure medications associated with more treatment of anxiety/depression?

**DOI:** 10.64898/2026.02.20.26344705

**Authors:** Meenakshi Singh, David Larsson, Johan Zelano

## Abstract

**Background:** Persons with epilepsy are at increased risk of depression/anxiety. Older antiseizure medications (ASMs) had drug-drug interactions that complicated pharmacotherapy of depression/anxiety; newer ASMs lack this drawback but can have psychiatric side effects. Anxiety/depression are increasingly recognized and treated pharmacologically. We hypothesized that the likelihood of treatment with selective serotonin uptake inhibitors (SSRI) would have increased in adult-onset epilepsy when prescription habits shifted towards newer ASMs.

**Methods:** We linked national health registers and included 28569 persons with epilepsy incident in 2006-2020 and 68509 age– and sex matched controls. We assessed the risk of starting SSRI treatment compared to age– and sex-matched controls across three incidence periods: 2006–2010, 2011–2015, and 2016–2020. Cox regression was used to estimate adjusted hazard ratios (HRs), and subgroup analyses explored age, sex, and comorbidities. Specialist psychiatric care was also assessed as a measure of more severe depression. Analysis including persons with SSRI-use before the epilepsy diagnosis were used for sensitivity analyses.

**Findings:** Persons with epilepsy had higher risks of starting SSRIs compared to controls; 1986/9561 (20.8%) received SSRI during follow-up after epilepsy in 2006-2010 and 2020/9165 (22.0%) in 2016-2020; adjusted HRs were 1.92 (95%CI:1.79 – 2.06) in 2006-2010, 1.84 (95%CI:1.72-1.97) in 2011-2015, and 1.81 (95%CI:1.69 – 1.94) in 2016-2020. Among individuals aged 18-30 years at their epilepsy diagnosis, the proportion receiving SSRIs remained the same between the first and last calendar periods (18.2%). Because of increased treatment of controls, the adjusted HRs of SSRI-treatment decreased from 2.33, (95% CI:1.96 – 2.78) to 1.63, (95% CI 1.39 to 1.91). The HR of specialist psychiatric care was not significantly different between the time periods. Most comorbidities were consistently associated with increased likelihood of SSRI treatment, whereas intellectual disability decreased the likelihood in some periods.

**Interpretation:** We found no evidence of overall increased SSRI initiation or psychiatric care after the shift to newer ASMs. Person with epilepsy remain more likely to receive SSRI treatment, but probably not to a level matching the higher prevalence of depression. Increased SSRI treatment of younger age adults has not been matched by increased treatment of young adults with epilepsy. This suggests a potentially widening treatment gap and a need for increased recognition of depression in young adults with epilepsy.

**Funding:** Swedish Research Council (2023-02816), Swedish state through the ALF-agreement (ALFGBG-1006343), Knut och Ragnvi Jacobsson foundation, Swedish Society for Medical Research (S18-0040), Swedish Society of medicine (SLS-881501), Epilepsifonden, Rune och Ulla Amlövs stiftelse.

## Introduction

Epilepsy affects over 50 million individuals and is commonly accompanied by psychiatric comorbidities, with depression being the most frequent.^1^ Depression affects quality of life and epilepsy treatment outcomes.^2^ The lifetime prevalence of depression in epilepsy is estimated at 30%^3,4^, and there is a bidirectional relationship between epilepsy and depression and anxiety.^5–7^ In addition to neurobiological links between the conditions, antiseizure medications (ASMs) used in epilepsy can have psychiatric side effects.^8^

In the last decades, there has been a shift in treatment of epilepsy from older ASMs like carbamazepine to newer ones like levetiracetam.^9^ Many older ASMs have drug-drug interactions making concomitant pharmacological treatment more difficult and perhaps discouraging such treatment in persons with epilepsy. Whether the shift to newer ASMs has increased treatment of comorbid depression in epilepsy is not known, but plausible given the increased recognition of psychiatric conditions in the last 15 years. We hypothesized that the use of SSRI – the most common treatment for depression and anxiety would have increased among epilepsy patients relative to the general population following the altered ASM prescription habits. We used Swedish national registers to examine the likelihood of SSRI treatment in the years following an epilepsy diagnosis between 2006 and 2020, with follow-up until 2023. We also studied specialized psychiatric care contacts as a measure of more severe psychiatric symptoms.

## Methods

### Registers

We identified the cohort from the Swedish Health Registers, particularly the National Patient Register (NPR) and the National Prescribed Drug Register (NPR). The data from these registers include diagnostic codes from inpatient care, outpatient specialist visits, emergency room visits, as well as all prescription drug dispensations at pharmacies across Sweden. Date of death was obtained from the Cause of Death Register (DR).

### Study population

Cases were identified by The National Board of Health and Welfare, using the following inclusion criteria: first diagnosis of epilepsy (ICD-10 code G40) recorded in the NPR as either a primary or secondary diagnosis after January 1, 2006, together with at least one prescription of an ASM (ATC code N03A) subsequent to epilepsy diagnosis. The combination of an epilepsy diagnosis with ASM strengthens the specificity of epilepsy identification within administrative data^10^. For each case, Statistics Sweden identified three controls without any epilepsy diagnosis from the general population register, matched for age and sex at the time of epilepsy diagnosis. The study population was categorized into three calendar periods based on the year of first epilepsy diagnosis or the index date for controls (2006-2010, 2011-2015, and 2016-2020). Exclusion criteria included: (1) inconsistent data, such as death before epilepsy diagnosis or first ASM retrieval, re-used personal ID numbers, and death of controls before the index date; (2) ASM (N03A) retrieval 3 months or more before diagnosis date of G40, G41, or R568 in cases (3) cases with any seizure-related diagnosis (G40, G41, R568) or ASMs (N03A) before January 1, 2006; (4) cases with any antidepressant medication (defined as ATC code N06A) before first epilepsy diagnosis or inclusion date; (5) controls with any seizure-related diagnosis (G40, G41, R568) or ASMs (N03A); (6) Seizure-related diagnosis (G40 or G41 or R568) or ASMs (N03A) before the age of 18. Inclusion and exclusion in number and percentage of cases/controls are stated in Suppl table 1.

### Outcomes

Time to new-onset SSRI treatment was defined as the time from the first ASM dispensation following the index epilepsy diagnosis (baseline) to the first dispensation of antidepressant medication (ATC code: N06A). Individuals were followed until the earliest of the following end-of-follow-up events: occurrence of the outcome (first antidepressant dispensation), death, emigration, or the end of the follow-up period for each calendar time frame – December 31, 2013, for the period 2006-2010; December 31, 2018, for the period 2011-2015; and December 31, 2023, for the period 2016-2020. First specialist psychiatric care was defined as occurrence of ICD-10 code F32-39 as main diagnosis.

### Demographics and Comorbidities

Information on age, sex, and comorbidities at the index date (first ASM dispensation) was obtained from the NPR. Comorbidities were defined as binary variables and included: stroke (ICD-10: I60–I64), traumatic brain injury (TBI; ICD-10: S06, S020, S021, S027, S029), dementia (ICD-10: F00–F03, G30, G31), brain tumor (ICD-10: C71, C793, D430, D32, D330), brain infection (ICD-10: A84, B004, A390, G00, G01, A17, A066, B431, G060, G079, A85, A86, B011, B020, B050, B262, B602, A87, B003, B010, B051, B261, B021, B375, B384, B582, G02), diabetes (ICD-10: E10, E11), cardiovascular diseases (ICD-10: I00–I99, excluding I10, I60–64, I11.9–I16), intellectual disability (ICD-10: F7), and cancer (ICD-10: C). The Charlson Comorbidity Index was calculated using a validated adaptation for Swedish registers.

### Statistical Analyses

Continuous variables were summarized using mean, standard deviation (SD), median, and range, while categorical variables were described using frequencies and percentages. Fisher’s exact test was used for binary comparisons, and Mann-Whitney U tests for continuous variables.

Cox regression was used to estimate the risk of new-onset SSRI-treatment among individuals with epilepsy compared to controls within each calendar period. Crude event rates were calculated as the number of events divided by follow-up time and expressed per 100 person-years. Exact Poisson limits were used to estimate 95% confidence intervals. To assess differences across calendar periods, an interaction term between period and group (case vs control) was included in the model.

Both unadjusted (matched for age and sex) and adjusted analyses (including age, sex, and comorbidities) were conducted for the overall population and all subgroups. Cox regression was also used to evaluate the association between baseline comorbidities and risk of new-onset depression in epilepsy cases, separately for each calendar period, and to assess changes in the importance of these variables over time. In cases where the month and day of diagnosis, medication, or death were missing, July 1 was used. In cases where the day of diagnosis, medication, or death was missing, day 15 was imputed. Otherwise, no other missing data was identified.

All statistical tests were two-tailed. A significant level of 0.05 was used for all analyses. All analyses were conducted using SAS software version 9.4 (SAS Institute Inc., Cary, NC, USA).

Sensitivity analyses were performed using cause-specific Cox regression to assess the robustness of the results.

### Subgroups

In addition to the overall population, analyses were conducted in predefined subgroups: based on the epilepsy onset (<30, 30–64, ≥65 years), stratified by sex.

### Ethical Permission and Data Handling

The study was approved by the Swedish Ethics Review Authority (approval number 2023-5598,17 Oct 2023), with waived need for informed consent. Data were linked by register holders using the unique Swedish personal identification number and de-identified prior to researcher access.

## Results

### Demographics

We included 28569 PWE incident in 2006-2020 and no prior SSRI use and 68509 age– and sex matched controls (eTable 1 in the supplement). The study population was stratified by calendar period for the first epilepsy diagnosis (2006-2010, 2011-2015, and 2016-2020) (Table 1 and eTable 2A – 2D in the supplement). The first ASM prescribed changed from carbamazepine being the most common in the first time period and levetiracetam being the most common in the last time period.) (eFigure 1A 1B, 1C, 1D in the supplement).

**Figure 1.**
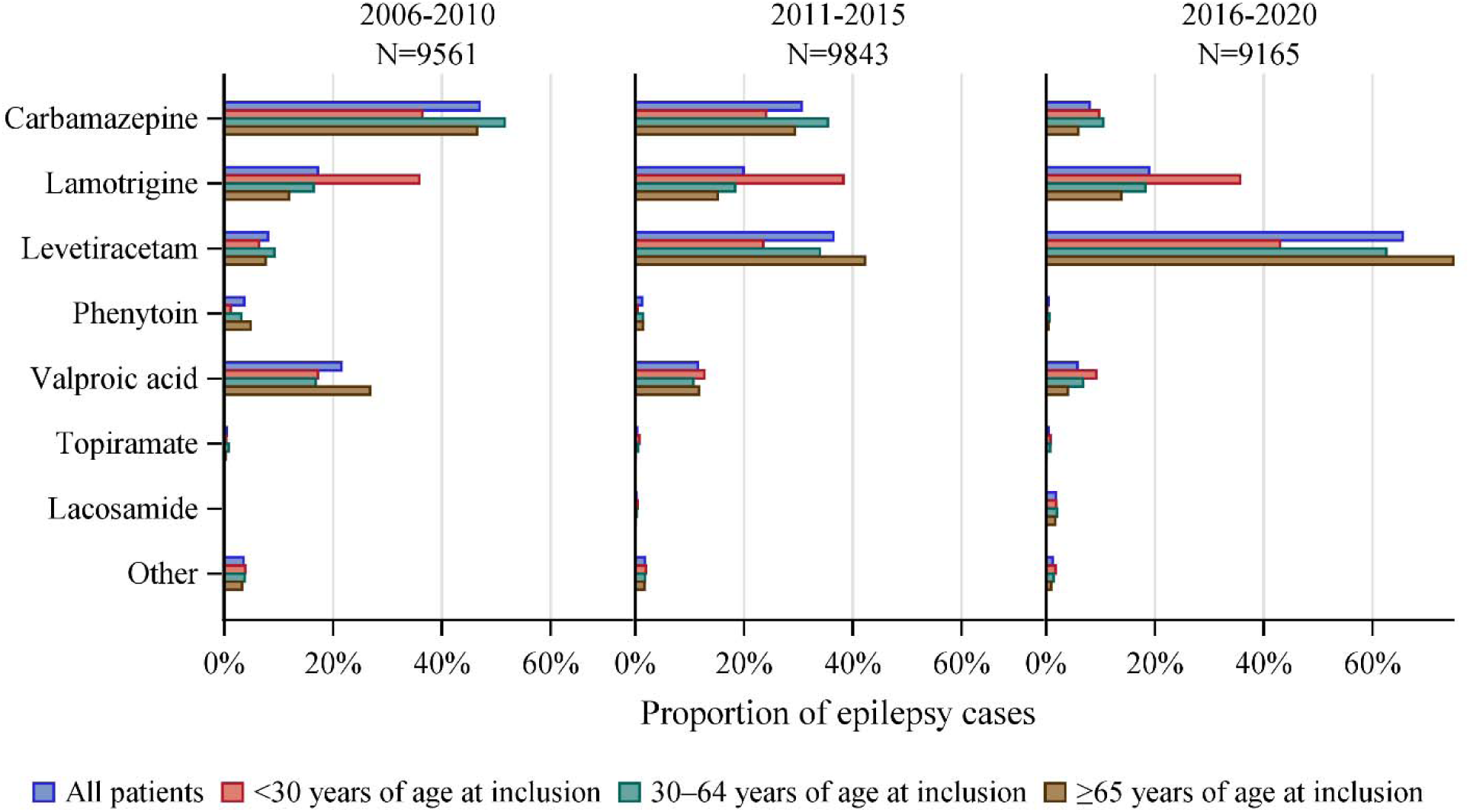
Shifts in first antiseizure medication across the temporal cohorts.

**Table 1.**
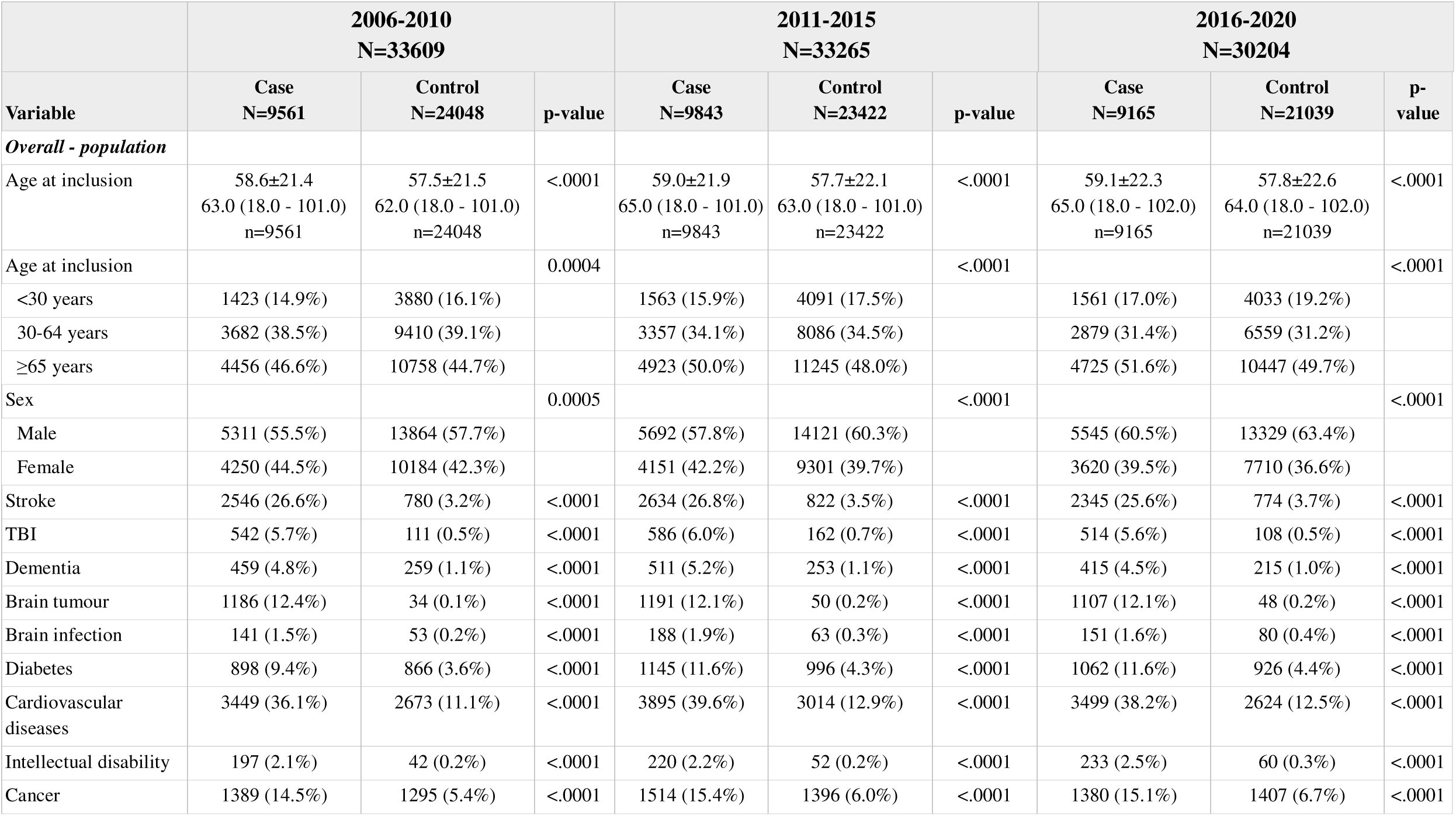
Baseline characteristics for cases and controls for calendar periods 2006-2010, 2011-2015, 2016-2020 overall and by sex.

### Trends in older and Newer ASMs

Carbamazepine decreased as the first ASM and levetiracetam increased from 2006 to 2020 (Figure 1A).

### Temporal trends in SSRI use

The proportion of patients starting SSRI after an epilepsy diagnosis in 2006-2010 was 20.8% vs. 11.0% in controls, resulting in a hazard ratio (aHRs) 1.92 (95% CI: 1.79 – 2.06). In 2011-2015, 21.1% of patients with epilepsy vs. 11.5% of controls (aHR 1.84, 95% CI: 1.72 – 1.97) and in 2016-2020, 22.0% of PWE and 12.2% for controls aHR 1.81 (95% CI: 1.69 – 1.94) (Figure 2A, eTable 3A. in the supplement). Overall, females had higher absolute rates of being prescribed SSRIs. Males exhibited slightly higher HRs relative to controls.

**Figure 2.**
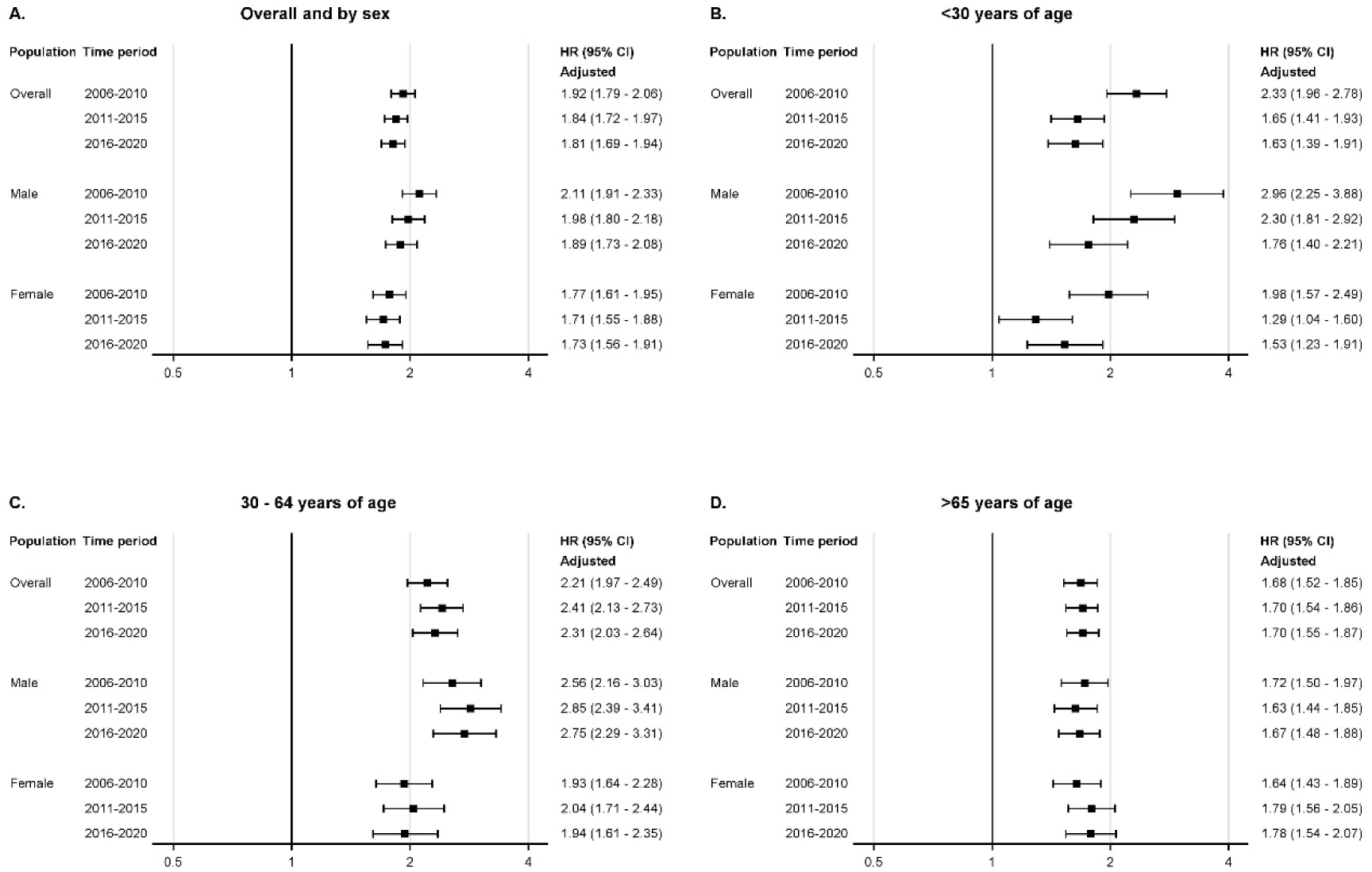
(**A**), Forest plot for fully adjusted HR, CIs, p-values for new onset depression overall and by sex. **(B),** For fully adjusted HR, CIs, p-values for new onset depression overall and by sex <30 years of age. **(C),** For fully adjusted HR, CIs, p-values for new onset depression overall and by sex 30 – 64 years of age. **(D),** For fully adjusted HR, CIs, p-values for new onset depression overall and by sex >65 years of age. Extreme left is population, followed by time period in years and extreme right is adjusted HR (95% CI)

In age-stratified analysis, individuals younger than 30 years (Figure 2B) at epilepsy onset had the highest HR, but lower absolute rates than older age groups. In the younger group, a total of 18.2% of person with epilepsy vs 8.1% in controls started SSRIs after inclusion in 2006-2010, (aHRs 2.33, 95% CI: 1.96 – 2.78). In 2016-2020 the proportion was still 18.2% for epilepsy cases but 11.1% in controls, resulting in an aHR of 1.63 (95% CI: 1.39 – 1.91) (Figure 2B, eTable 3B. in the supplement).

In age groups the 30-64 years (Figure 2C, (eTable 3B. in the supplement), the risk relative controls remained elevated and the highest among all groups. The absolute percentage for the events in 2006-2010 was (19.7%) for epilepsy cases vs. (8.6%) in controls (aHR 2.21, 95% CI: 1.97 – 2.49). In 2011-2015 is (20.6%) for epilepsy cases vs. (8.0%) in controls with adjusted HRs 2.41 (95% CI: 2.13 – 2.73). In 2016-2020 is (21.3%) for epilepsy cases vs. (8.7%) in controls with aHRs 2.31 (95% CI: 2.03 – 2.64) across the three time periods, respectively.

Individuals aged 65 years and older (Figure 2D, eTable 3B. in the supplement had the highest likelihood of starting SSRIs.

For those diagnosed in 2006-2010, 22.5% of person with epilepsy compared to 14.2% in controls started SSRI (aHR 1.68 95% CI: 1.52 – 1.85),). In 2016-2020 is (23.8%) for epilepsy cases vs. (14.8%) in controls with adjusted HRs 1.70 (95% CI: 1.55 – 1.87. Unadjusted values corresponding to Figure 2A–D are provided in the supplement (eFigure 2A–D).

### Specialist Psychiatric Care

The need for specialist psychiatric care was also significantly higher among PWE and did not change across age groups and calendar periods.

In the overall population (Figure 3A, eTable 4A. in the supplement), the absolute percentage for the events in 2006-2010 is (2.4%) for cases vs. (0.9%) in controls with unadjusted HRs for psychiatric care were 3.35 (95% CI: 2.41 – 3.66). In 2011-2015 it is (2.1%) for cases vs. (0.8%) in controls with unadjusted HRs 2.95 (95% CI: 2.16 – 3.32). In 2016-2020 is (1.6%) for cases vs. (0.5%) in controls with unadjusted HRs 3.87 (95% CI: 2.69 – 4.65) all with p-values < 0.0001.

**Figure 3.**
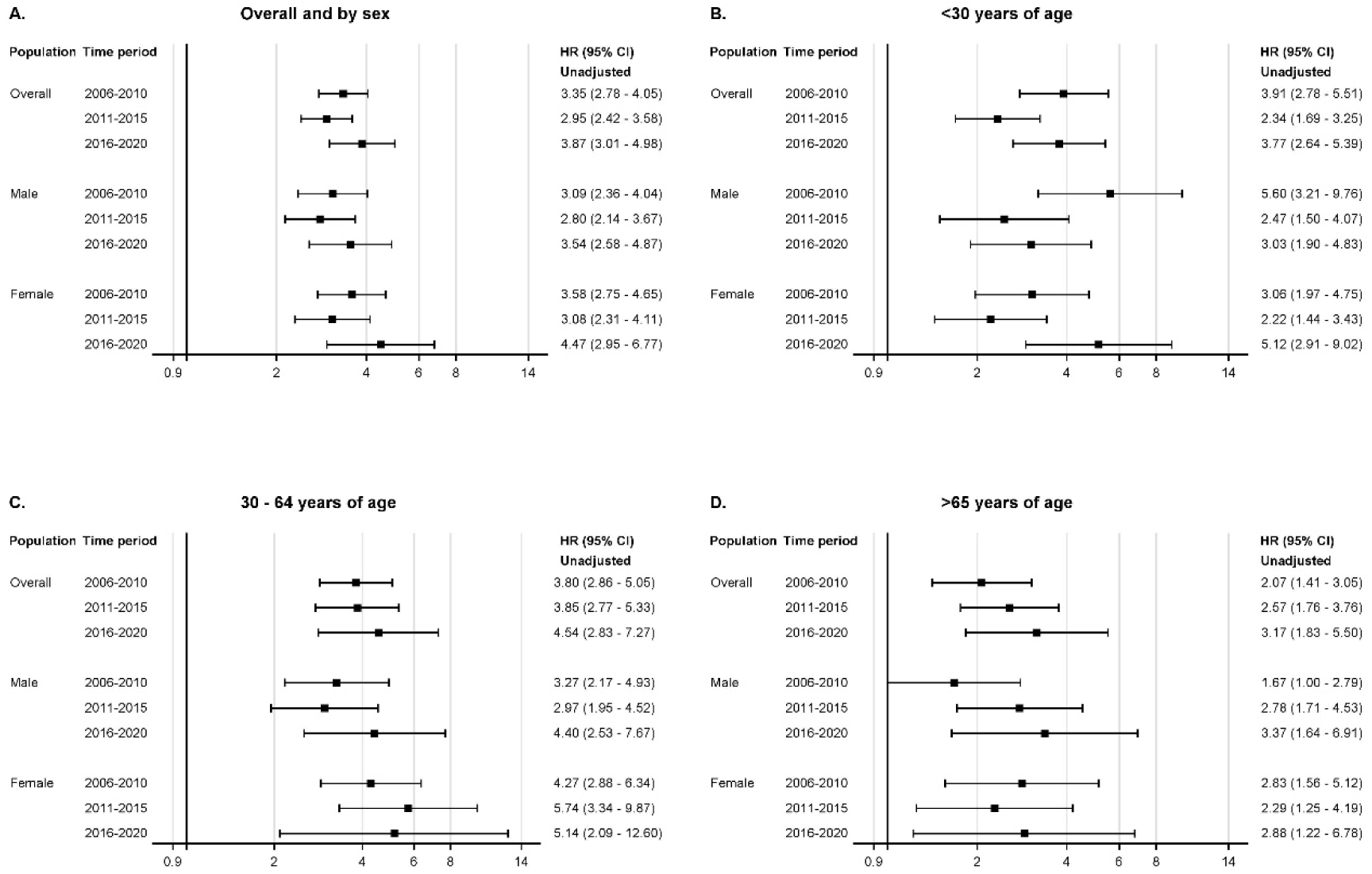
(**A**), Forest plot for unadjusted HR, CIs, p-values for first specialist psychiatric care overall and by sex. **(B),** For unadjusted HR, CIs, p-values for first specialist psychiatric care overall and by sex <30 years of age. **(C),** For unadjusted HR, CIs, p-values for first specialist psychiatric care overall and by sex 30-64 years of age. **(D),** For unadjusted HR, CIs, p-values for for first specialist psychiatric care overall and by sex 65 years of age. Extreme left is population, followed by time period in years and extreme right is adjusted HR (95% CI)

Among those under 30 years (Figure 3B eTable 4B. in the supplement), the relative risk was particularly pronounced.

In the 30-64 years group (Figure 3C), eTable 4B. in the supplement), the absolute percentage for the events in 2006-2010 is (3.0%) for cases vs. (0.9%) in controls with the unadjusted HRs were 3.80 (95% CI: 2.86 – 5.05). In 2011-2015 is (2.6%) for cases vs. (0.8%) in controls with unadjusted HRs 3.85 (95% CI: 2.77 – 5.33). In 2016-2020 is (1.7%) for cases vs. (0.4%) in controls with unadjusted HRs 4.54 (95% CI: 2.83 – 7.27).

For the ≥65 years group (Figure 3D, eTable 4B. in the supplement) although the relative risk was slightly lower, it remained significant with the absolute percentage for the events. In 2006-2010 the absolute percentage for the events is (1.0%) for cases vs. (0.6%) in controls with unadjusted HRs of 2.07 (95% CI: 1.41 – 3.05). In 2011-2015 is (1.0%) for cases vs. (0.5%) in controls with unadjusted HRs 2.57 (95% CI: 1.76 – 3.76). In 2016-2020 is (0.6%) for cases vs. (0.2%) in controls with unadjusted HRs 3.17 (95% CI: 1.83 – 5.50). Adjusted values corresponding to Figure 3A are provided in the supplement (eFigure 3A).

Cumulative incidence curves for new-onset depression and specialist psychiatric care are shown in eFigure 4A and B in the Supplement).

### Comorbidity associated with starting SSRIs

Several comorbid conditions were predictors of SSRI treatment among PWE. In different time periods the stroke was associated with an HR ranging 1.94 (95% CI: 1.77 – 2.12, in the calendar period 2006-2010), 1.65 (95% CI: 1.50 – 1.81, in 2016-2020), and the interaction p-value between the calendar period is 0.012, while TBI showed a more modest but still significant association with an HR ranging 1.20 (95% CI: 1.01 – 1.44, in the calendar period 2006-2010), 1.33 (95% CI: 1.12 – 1.59, in 2016-2020) and the p-value for the interaction between the calendar period is 0.43. Dementia with an HR ranging 1.38 (95% CI: 1.11 – 1.70, in the calendar period 2006-2010), 1.80 (95% CI: 1.48 – 2.20, in 2016-2020) and the p-value for the interaction between the calendar period is 0.09. Brain tumors with an HR ranging 1.32 (95% CI: 1.14 – 1.52, in the calendar period 2006-2010), 1.58 (95% CI: 1.38 – 1.80, in 2016-2020) and the p-value for the interaction between the calendar period is 0.09. Diabetes with an HR ranging 1.30 (95% CI: 1.13 – 1.51, in the calendar period 2006-2010), 1.31 (95% CI: 1.15 – 1.50, in 2016-2020) and the p-value for the interaction between the calendar period is 0.95. Cardiovascular disease with an HR ranging 1.55 (95% CI: 1.42 – 1.70, in the calendar period 2006-2010), 1.51 (95% CI: 1.38 –-1.65, in 2016-2020) and the p-value for the interaction between the calendar period is 0.64. Cancer with an HR ranging 1.37 (95% CI: 1.20 – 1.56, in the calendar period 2006-2010), 1.66 (95% CI: 1.47 – 1.87, in 2016-2020) and the p-value for the interaction between the calendar period is 0.053, were all independently associated with an increased risk of depression. Intellectual disability had HRs below 1 (eTable 5. in the supplement). For instance, during 2006-2010, the HR for depression among individuals with intellectual disability was (0.60, 95% CI: 0.41 – 0.86 and the p-value for the interaction between the calendar period is 0.78., (eTable 5. in the supplement).

### Sensitivity analysis

To ensure that the results were not due to exclusion of prior SSRI use before the diagnosis of epilepsy, perhaps reflecting psychiatric vulnerability, we also performed a sensitivity analysis in which patients with SSRI use before the epilepsy were not excluded. This analysis included 45,579 PWE and 130,557 controls and yielded results similar to those of the main analysis. using SSRI after an epilepsy diagnosis in 2006-2010 was 2.85 (95% CI: 2.51 – 3.24, 29.4% of persons with epilepsy vs 11.2% of controls) and had decreased to 2.06 (95% CI: 1.86 – 2.28) in 2016-2020 (33.2% for epilepsy cases and 17.7% in controls) largely due to the increase in treatment of controls. In contrast, there was no significant decrease over the time periods for those aged 30-65; SSRI use after epilepsy diagnosed in 2006-2010 was seen in 39.4% of epilepsy vs. 16.0% in controls (aHR 2.64, 95% CI: 2.47 – 2.83) and after epilepsy diagnosed in 2016-2020 in 43.0% vs. 19.5% in controls (aHR 2.33, 95% CI: 2.19 – 2.49). In those aged 65 and older, subsequent SSRI use was detected in 41.9% and 24.9% in person with epilepsy diagnosed in 2006-2010 vs controls, respectively (aHR 1.53 95% CI: 1.45 – 1.62), and the aHR was unchanged in 2016-2020; (44.3% vs 28.2%, aHR 1.54, 95% CI: 1.47 – 1.62).

## Discussion

We found no overall increase in the use of antidepressants or psychiatric in or outpatient care among patients with new-onset epilepsy over 2006-2020, with follow-up through 2023. This suggests that despite newer ASMs being less complicated from a drug-drug interaction perspective and an increased recognition of psychiatric symptoms in society in general, depression or anxiety in epilepsy are not treated to any greater extent.

PWE had an approximately two-fold likelihood of starting SSRIs or seeking psychiatric care in the years following the epilepsy diagnosis. The observed hazard ratios for starting SSRIs, ranging from 1.81 to 1.92 across the three calendar periods, are in line with previous observations of risks of depression in epilepsy.^11,12^ An interesting observation is that the controls of the youngest (<30) age group became increasingly likely to receive SSRIs, which was not seen in persons of the same age with epilepsy. It seems that increased recognition of depression/anxiety in younger patients has not extended to person with epilepsy.

We also studied utilization of specialized psychiatric care as an indicator of more severe psychiatric symptoms. The increased risk of requiring specialist psychiatric care, with hazard ratios ranging from 2.68 to 3.54 did not show any marked changes over the study time periods. Notably, the highest HR was observed in the most recent period (2016–2020), which may reflect improved detection and referral practices, greater patient awareness, or a true increase in severe psychiatric morbidity.

Several comorbid conditions were identified as predictors of SSRI use in epilepsy, including stroke, traumatic brain injury, dementia, brain tumors, diabetes, cardiovascular disease, and cancer. These findings are consistent with the broader literature on multimorbidity and mental health.^1,13^ The strong associations observed with neurological comorbidities such as stroke and dementia may reflect guidelines emphasizing recognition of clinical symptoms.^5^

A somewhat unexpected finding was the low likelihood of person with epilepsy and intellectual disability to receive SSRI treatment. This could indicate that depression is either unrecognized or remains untreated in patients with intellectual disability due to communication barriers or diagnostic overshadowing. Social support structures or care environments could also play a role.^14^ Further qualitative and quantitative research is needed to examine this association in more depth and determine whether interventions effective in the general population are equally effective here.

From a clinical perspective, this highlights the importance of developing specialized screening tools and assessment strategies that are required for the unique presentation of mood disorders in people with intellectual disability. A missed diagnosis could lead to poorer long-term outcomes.

There has been a discussion that levetiracetam, the most popular new ASM, has greater potential for psychiatric side effects.^15–17^ In our analysis, the shift to levetiracetam has not led to more treated depression or psychiatric care in any age group.

Taken together, our findings indicate that recognition and treatment of depression in new onset of epilepsy in the last 20 years in Sweden. Routine screening for depression and other psychiatric conditions in combination with multidisciplinary care models could be an option.^18,19,20^

## Conclusion

This study highlights consistently elevated risk of new-onset depression, psychiatric care and SSRI initiation among individuals with epilepsy, with hazard ratios nearing two-fold compared to controls. Age-specific analyses revealed that younger adults face the highest relative risks, while older adults bear the greatest absolute burden. Several comorbid conditions, particularly neurological and chronic physical illnesses, were significant predictors of depression.

### Authors List

First Author – Meenakshi Singh^1,3^, Corresponding author – Meenakshi Singh^1,3^ and Johan Zelano^1,3^

### Additional Contribution

We thank Christopher Backström (APNC AB, Gothenburg, Sweden) for his expert assistance with the statistical analyses of the data.

Role of the Funding Source

The authorized funding sources pursue no role in the study design; data collection, data analysis, or the interpretation of the data; neither in writing the report; nor in the decision to submit the manuscript for publication.

### Conflict of Interest/Ethical Publication Statement

JZ reports speaker / advisory board honoraria from UCB, Eisai, Orion Pharma, Angelini Pharma, and Sanofi and as an employee of Sahlgrenska University Hospital being investigator in trials sponsored by SK life science, Bial, UCB, GW Pharma, and Angelini Pharma (no personal compensation).

DL reports no conflicts of interest.

The funding sources or institute have had no influence on any part of this study or decision to publish.

## Supporting information

.docx

.docx

## Data Availability

The underlying register data cannot be shared by the authors due to Swedish privacy regulations but are available from the register holders following application.

## Notes

### Funding Statement

The study eas funded by Swedish Research Council (2023–02816), Swedish state through the ALF-agreement (ALFGBG –1006343), Knut och Ragnvi Jacobsson foundation, Swedish Society for Medical Research (S18–0040), Swedish Society of medicine (SLS –881501), Epilepsifonden, Rune och Ulla Amlövs stiftelse

### Author Declarations

Ethical Permission and Data Handling The study was approved by the Swedish Ethics Review Authority (approval number 2023–5598, 17 Oct 2023), with waived need for informed consent. All data were linked by register holders using the unique Swedish personal identification number and anonymized prior to researcher access.

