## Supplementary material for "Incidence of SSRI treatment and psychiatric specialist care in new-onset adult epilepsy: are newer antiseizure medications associated with more treatment of anxiety/depression?": .docx

**Supplementary eFigures Online Content**

**eFigure 1A.** Bar graph, proportion of epilepsy cases receiving different ASMs as the first ASM after diagnosis of epilepsy per calendar periods, overall and by sex (FAS population)

**eFigure 1B.** Bar graph, proportion of epilepsy cases receiving different ASMs as the first ASM after diagnosis of epilepsy per calendar periods, overall and by sex (FAS population - older patients)

**eFigure 1C.** Bar graph, proportion of epilepsy cases receiving different ASMs as the first ASM after diagnosis of epilepsy per calendar periods, overall and by sex (FAS population - Patients with cardiovascular diseases or stroke)

**eFigure 1D.** Bar graph, proportion of epilepsy cases receiving different ASMs as the first ASM after diagnosis of epilepsy per calendar periods, overall and by sex (FAS population - Patients with generalized epilepsy)

**eFigure 2A.** Forest plot for unadjusted, and fully adjusted HR, CIs, p-values for new-onset depression overall and by sex (FAS population)

**eFigure 2B**. Forest plot for unadjusted, and fully adjusted HR, CIs, p-values for new-onset depression overall and by sex (FAS population - <30 years of age)

**eFigure 2C.** Forest plot for unadjusted, and fully adjusted HR, CIs, p-values for new-onset depression overall and by sex (FAS population - 30-64 years of age)

**eFigure 2D.** Forest plot for unadjusted, and fully adjusted HR, CIs, p-values for new-onset depression overall and by sex (FAS population - ≥65 years of age)

**eFigure 3A.** Forest plot for unadjusted, and fully adjusted HR, CIs, p-values for first specialist psychiatric care overall and by sex (FAS population)

**eFigure 4A.** Cumulative incidence curve for new-onset depression adjusted for death as competing risk for cases and controls (FAS population)

**eFigure 4B.** Cumulative incidence curve for specialist psychiatric care adjusted for death as competing risk for cases and controls (FAS population)

### **eFigure 1A.** Bar graph, proportion of epilepsy cases receiving different ASMs as the first ASM after diagnosis of epilepsy per calendar periods, overall and by sex (FAS population).

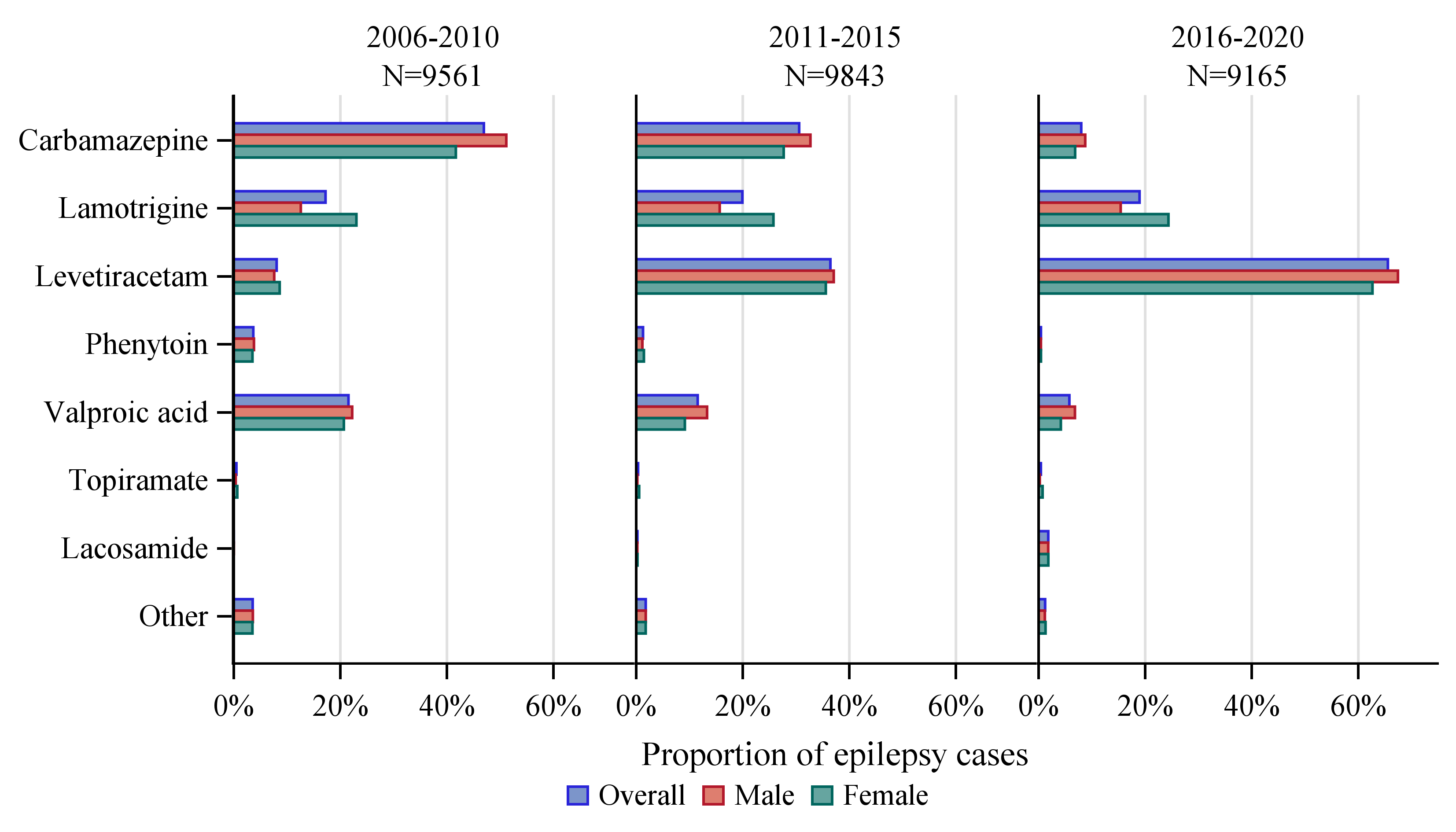

### **eFigure 1B.** Bar graph, proportion of epilepsy cases receiving different ASMs as the first ASM after diagnosis of epilepsy per calendar periods, overall and by sex (FAS population - older patients).

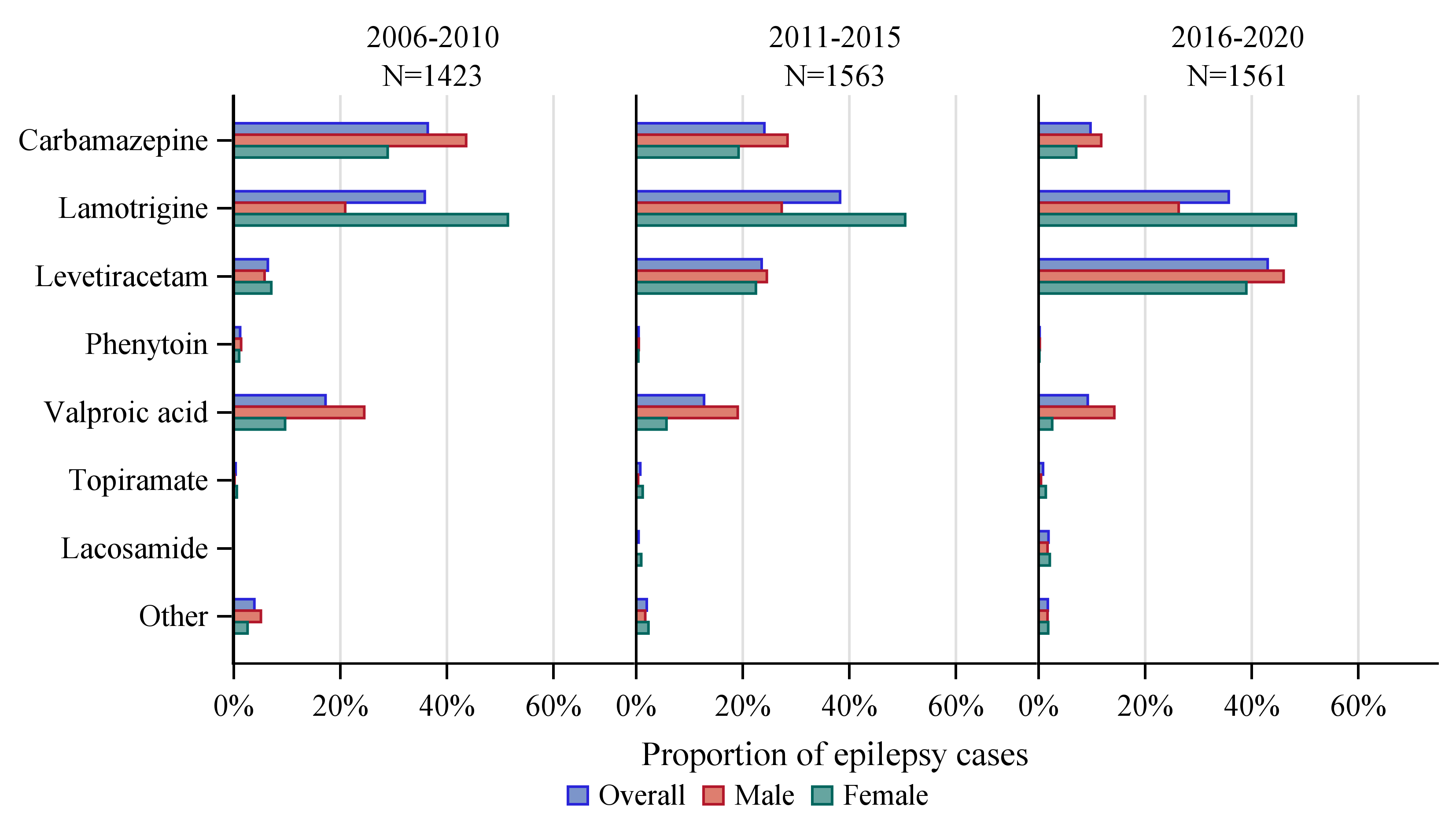

### **eFigure 1C.** Bar graph, proportion of epilepsy cases receiving different ASMs as the first ASM after diagnosis of epilepsy per calendar periods, overall and by sex (FAS population - Patients with cardiovascular diseases or stroke).

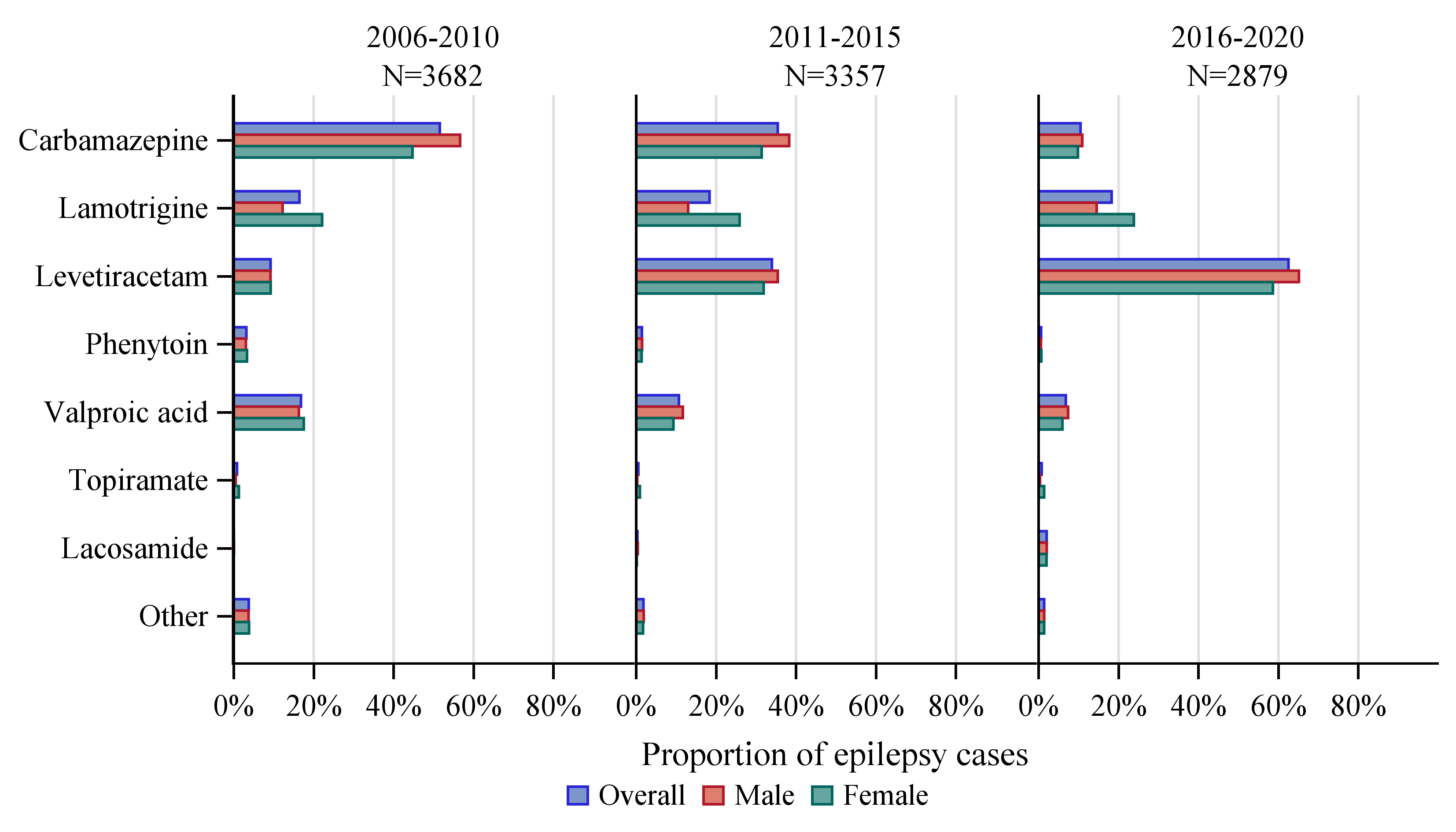

### **eFigure 1D.** Bar graph, proportion of epilepsy cases receiving different ASMs as the first ASM after diagnosis of epilepsy per calendar periods, overall and by sex (FAS population - Patients with generalized epilepsy).

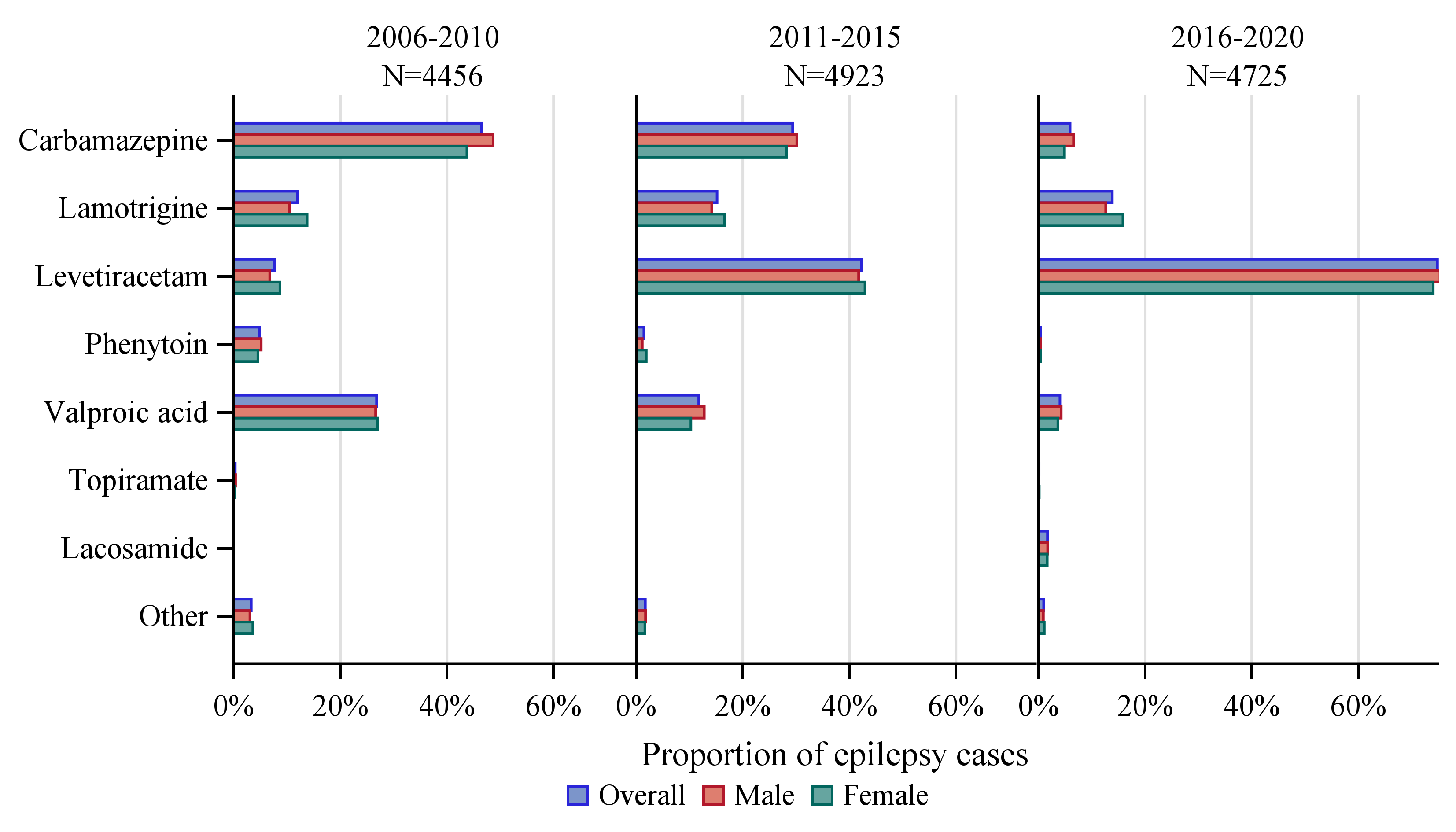

### **eFigure 2A.** Forest plot for unadjusted, and fully adjusted HR, CIs, p-values for new-onset depression overall and by sex (FAS population).

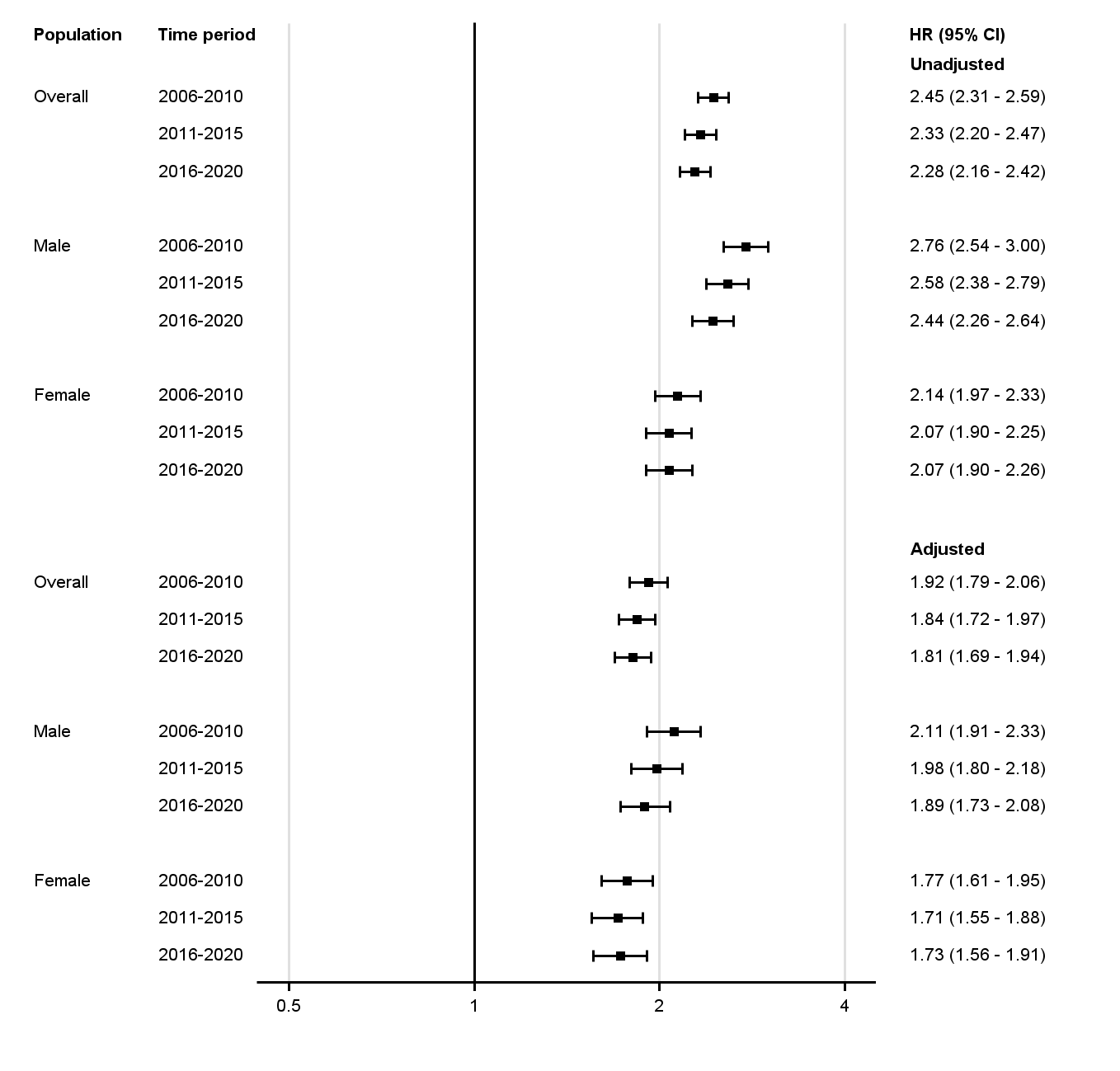

### **eFigure 2B.** Forest plot for unadjusted, and fully adjusted HR, CIs, p-values for new-onset depression overall and by sex (FAS population **-** <30 years of age).

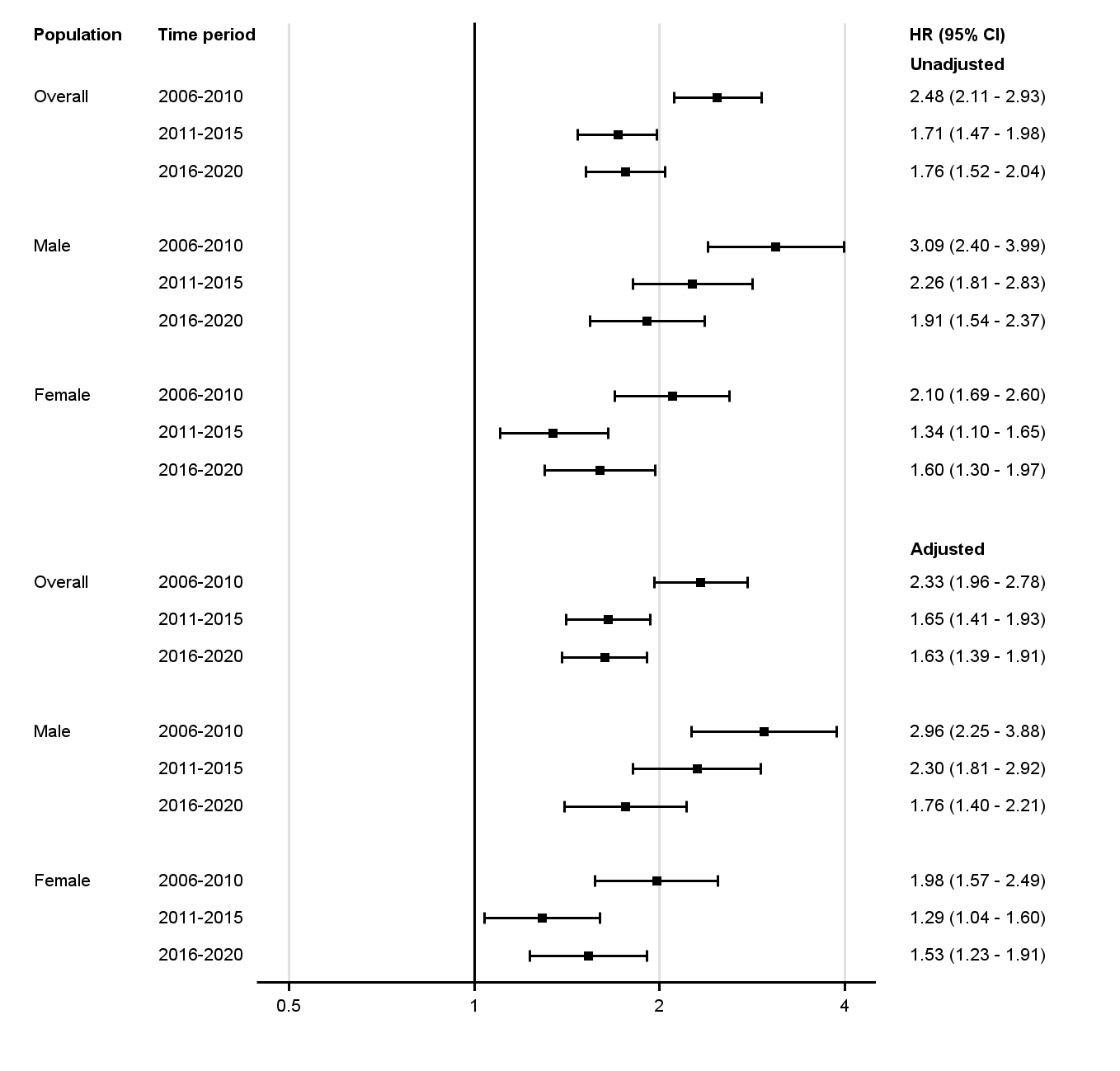

### **eFigure 2C.** Forest plot for unadjusted, and fully adjusted HR, CIs, p-values for new-onset depression overall and by sex (FAS population **-** 30-64 years of age).

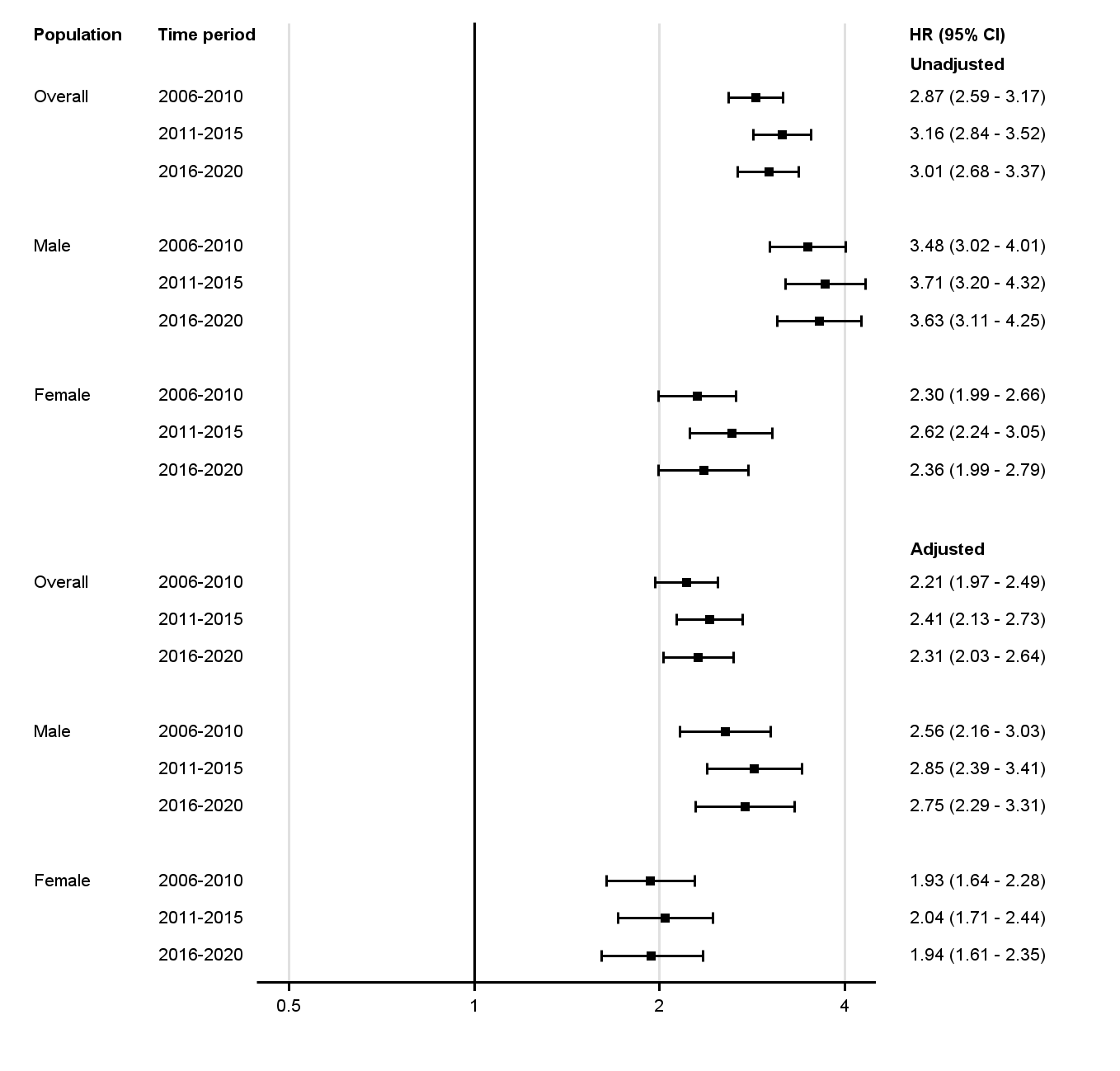

### **eFigure 2D.** Forest plot for unadjusted, and fully adjusted HR, CIs, p-values for new-onset depression overall and by sex (FAS population **-** ≥65 years of age).

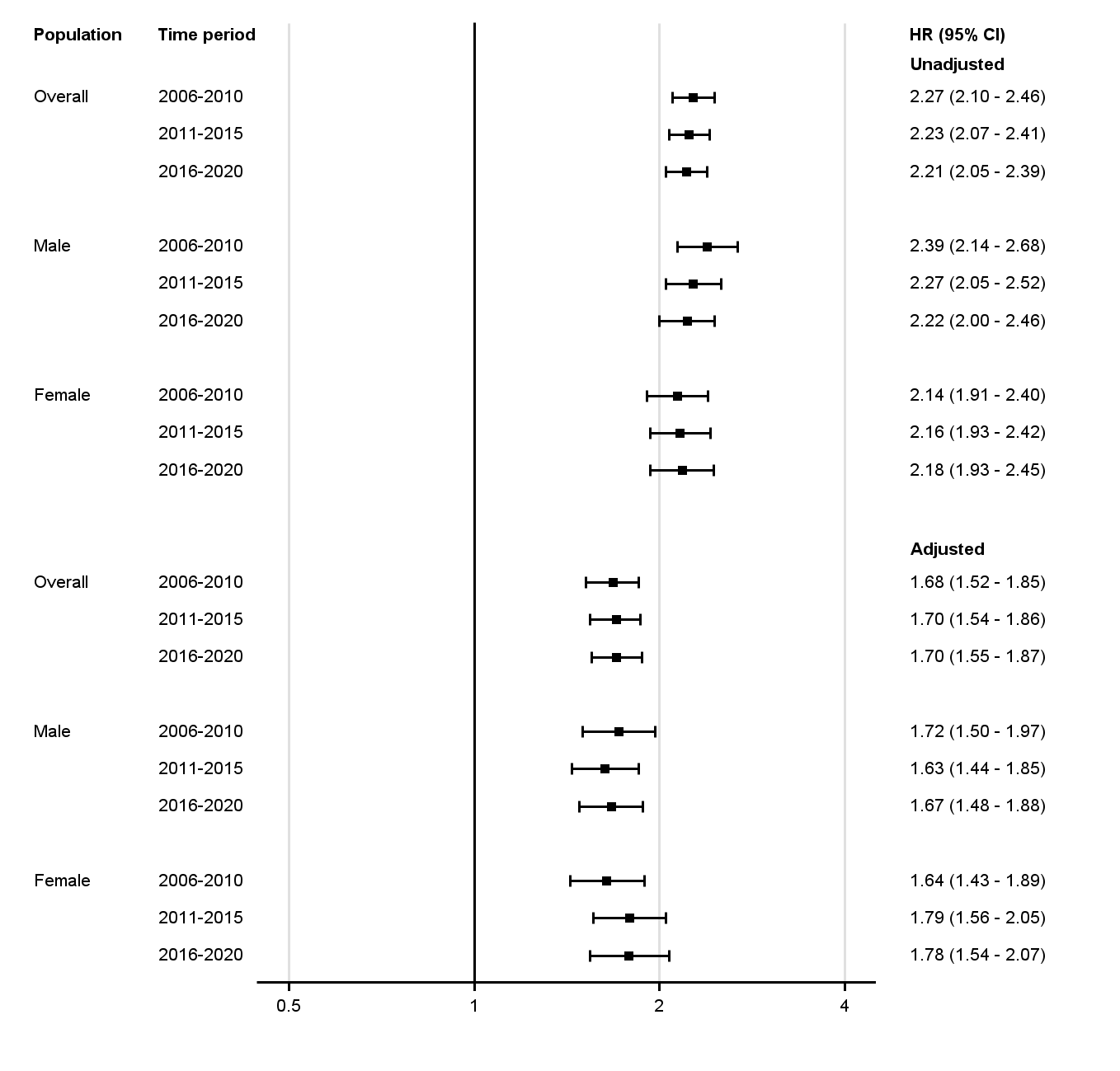

### **eFigure 3A.** Forest plot for unadjusted, and fully adjusted HR, CIs, p-values for first specialist psychiatric care overall and by sex (FAS population).

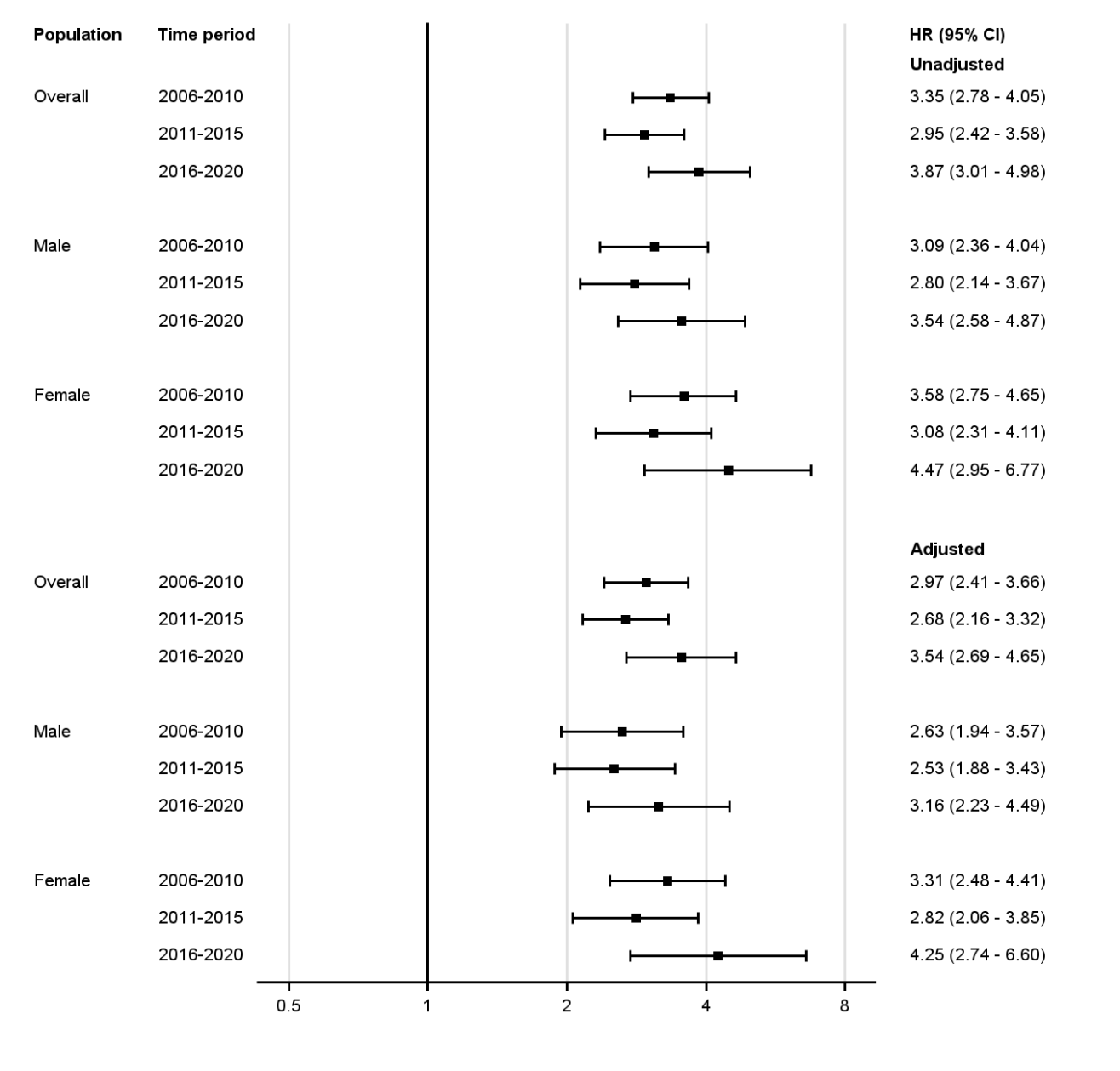

### **eFigure 4A.** Cumulative incidence curve for new-onset depression adjusted for death as competing risk for cases and controls (FAS population).

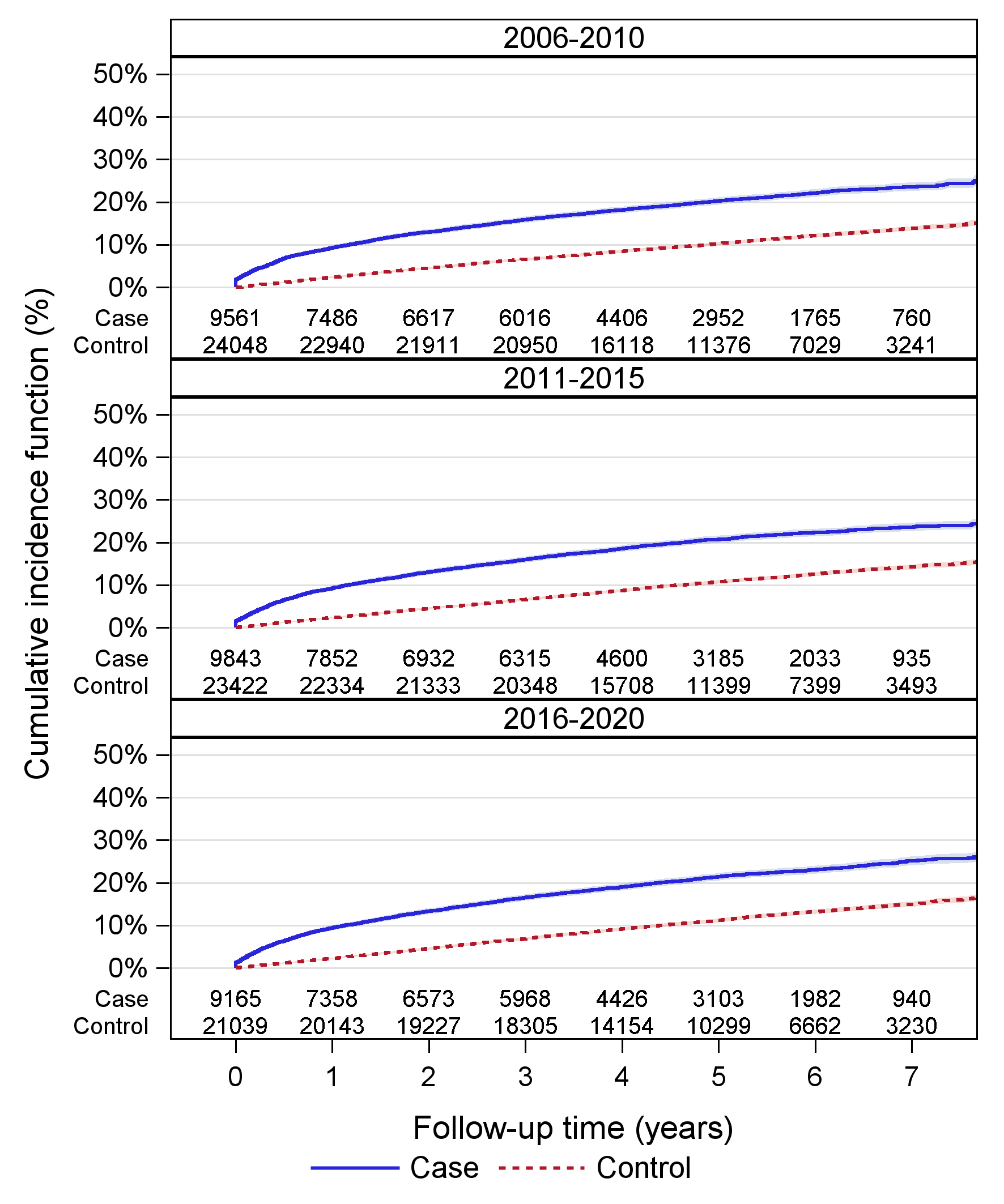

### **eFigure 4B.** Cumulative incidence curve for specialist psychiatric care adjusted for death as competing risk for cases and controls (FAS population).

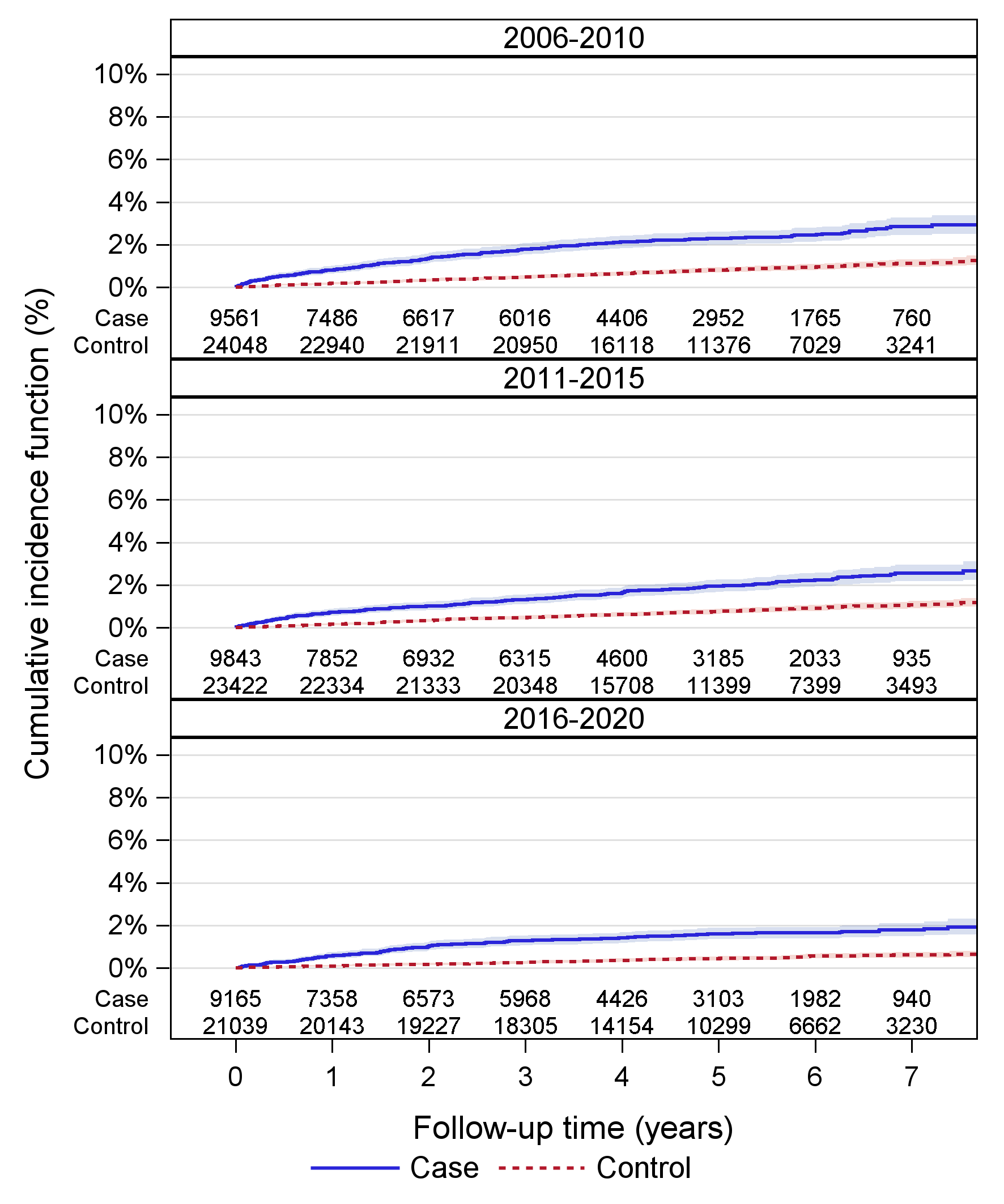
