## Supplementary material for "Incidence of SSRI treatment and psychiatric specialist care in new-onset adult epilepsy: are newer antiseizure medications associated with more treatment of anxiety/depression?": .docx

**Supplementary eTables Online Content**

**eTable 1.** Inclusions/exclusions

**eTable 2A**. Baseline characteristics for cases and controls, overall and for calendar periods 2006-2010, 2011-2015, 2016-2020 overall and by sex (FAS population)

**eTable 2B.** Baseline characteristics for cases and controls, overall and for calendar periods 2006-2010, 2011-2015, 2016-2020 overall and by sex (FAS population - Age at epilepsy onset <30 years)

**eTable 2C.** Baseline characteristics for cases and controls, overall and for calendar periods 2006-2010, 2011-2015, 2016-2020 overall and by sex (FAS population - Age at epilepsy onset 30-64 years)

**eTable 2D.** Baseline characteristics for cases and controls, overall and for calendar periods 2006-2010, 2011-2015, 2016-2020 overall and by sex (FAS population - Age at epilepsy onset ≥65 years)

**eTable 3A.** Unadjusted, and fully adjusted HR, CIs, p-values for new-onset depression from Cox regression in cases vs controls during periods 2006-2010, 2011-2015 and 2016-2020 overall and by sex (FAS population)

**eTable 3B.** Unadjusted, and fully adjusted HR, CIs, p-values for new-onset depression from Cox regression in cases vs controls during periods 2006-2010, 2011-2015 and 2016-2020 for each subgroup overall and by sex (FAS population)

**eTable 4A.** Unadjusted, and fully adjusted HR, CIs, p-values for first specialist psychiatric care from Cox regression in cases vs controls during periods 2006-2010, 2011-2015 and 2016-2020 overall and by sex (FAS population)

**eTable 4B.** Unadjusted, and fully adjusted HR, CIs, p-values for first specialist psychiatric care from Cox regression in cases vs controls during periods 2006-2010, 2011-2015 and 2016-2020 for each subgroup overall and by sex (FAS population)

**eTable 5.** Risk factors for new-onset depression from Cox regression during periods 2006-2010, 2011-2015 and 2016-2020 (FAS population - Epilepsy cases)

### **eTable 1 Inclusions/exclusions**

| **Included/Excluded** | **Total N=620244** |
| --- | --- |
| Number of patients included/excluded |  |
| Included | 97078 (15.7%) |
| Exclusion - Reusable personal identification number | 209 (0.0%) |
| Exclusion of case - Inconsistent data: Death date before first inclusion date | 79 (0.0%) |
| Exclusion of case - Inconsistent data: Missing date of epilepsy diagnosis | 2 (0.0%) |
| Exclusion of case - Inconsistent data: no ASM retrieval | 260 (0.0%) |
| Exclusion of case - Date of first epilepsy before 01JAN2006 | 42619 (6.9%) |
| Exclusion of case - Date of first seizure diagnosis before 01JAN2006 | 3856 (0.6%) |
| Exclusion of case - Date of first ASM before 01JAN2006 | 10597 (1.7%) |
| Exclusion of case - Date of first epilepsy diagnosis after 31DEC2020 | 16905 (2.7%) |
| Exclusion of case - All three controls matched to the respective case have been excluded | 383 (0.1%) |
| Exclusion of case - Date of inclusion after 31DEC2020 | 866 (0.1%) |
| Exclusion of case - ASM retrieval 3 months before first epilepsy diagnosis | 17864 (2.9%) |
| Exclusion of case - ASM used before first diagnosis of epilepsy but not after | 497 (0.1%) |
| Exclusion of control - Control with epilepsy diagnosis | 367 (0.1%) |
| Exclusion of control - Control with seizure diagnosis | 1035 (0.2%) |
| Exclusion of control - Control with ASM retrieval | 2138 (0.3%) |
| Exclusion of control - Control with death date before inclusion date of respective case | 255 (0.0%) |
| Exclusion of case - Seizure-related diagnosis (G40 or G41 or R568) or ASMs (N03A) before the age of 18 | 15796 (2.5%) |
| Exclusion of case/control - Antidepressive medication before first epilepsy diagnosis or inclusion date. | 31111 (5.0%) |
| Exclusion of control - Control excluded because respective case was excluded | 378327 (61.0%) |
| Data are presented as number (percentage). For each patient, only the first matching exclusion reason is assigned, in the order the reasons appear in the table from top to bottom. | |

### **eTable 2A. Baseline characteristics for cases and controls, overall and for calendar periods 2006-2010, 2011-2015, 2016-2020 overall and by sex (FAS population)**

|  | **2006-2010 N=33609** | | | **2011-2015 N=33265** | | | **2016-2020 N=30204** | | |
| --- | --- | --- | --- | --- | --- | --- | --- | --- | --- |
| **Variable** | **Case N=9561** | **Control N=24048** | **p-value** | **Case N=9843** | **Control N=23422** | **p-value** | **Case N=9165** | **Control N=21039** | **p-value** |
| ***Overall - population*** |  |  |  |  |  |  |  |  |  |
| Age at inclusion | 58.6±21.4 63.0 (18.0 - 101.0) n=9561 | 57.5±21.5 62.0 (18.0 - 101.0) n=24048 | <.0001 | 59.0±21.9 65.0 (18.0 - 101.0) n=9843 | 57.7±22.1 63.0 (18.0 - 101.0) n=23422 | <.0001 | 59.1±22.3 65.0 (18.0 - 102.0) n=9165 | 57.8±22.6 64.0 (18.0 - 102.0) n=21039 | <.0001 |
| Age at inclusion |  |  | 0.0004 |  |  | <.0001 |  |  | <.0001 |
| <30 years | 1423 (14.9%) | 3880 (16.1%) |  | 1563 (15.9%) | 4091 (17.5%) |  | 1561 (17.0%) | 4033 (19.2%) |  |
| 30-64 years | 3682 (38.5%) | 9410 (39.1%) |  | 3357 (34.1%) | 8086 (34.5%) |  | 2879 (31.4%) | 6559 (31.2%) |  |
| ≥65 years | 4456 (46.6%) | 10758 (44.7%) |  | 4923 (50.0%) | 11245 (48.0%) |  | 4725 (51.6%) | 10447 (49.7%) |  |
| Sex |  |  | 0.0005 |  |  | <.0001 |  |  | <.0001 |
| Male | 5311 (55.5%) | 13864 (57.7%) |  | 5692 (57.8%) | 14121 (60.3%) |  | 5545 (60.5%) | 13329 (63.4%) |  |
| Female | 4250 (44.5%) | 10184 (42.3%) |  | 4151 (42.2%) | 9301 (39.7%) |  | 3620 (39.5%) | 7710 (36.6%) |  |
| Stroke | 2546 (26.6%) | 780 (3.2%) | <.0001 | 2634 (26.8%) | 822 (3.5%) | <.0001 | 2345 (25.6%) | 774 (3.7%) | <.0001 |
| TBI | 542 (5.7%) | 111 (0.5%) | <.0001 | 586 (6.0%) | 162 (0.7%) | <.0001 | 514 (5.6%) | 108 (0.5%) | <.0001 |
| Dementia | 459 (4.8%) | 259 (1.1%) | <.0001 | 511 (5.2%) | 253 (1.1%) | <.0001 | 415 (4.5%) | 215 (1.0%) | <.0001 |
| Brain tumour | 1186 (12.4%) | 34 (0.1%) | <.0001 | 1191 (12.1%) | 50 (0.2%) | <.0001 | 1107 (12.1%) | 48 (0.2%) | <.0001 |
| Brain infection | 141 (1.5%) | 53 (0.2%) | <.0001 | 188 (1.9%) | 63 (0.3%) | <.0001 | 151 (1.6%) | 80 (0.4%) | <.0001 |
| Diabetes | 898 (9.4%) | 866 (3.6%) | <.0001 | 1145 (11.6%) | 996 (4.3%) | <.0001 | 1062 (11.6%) | 926 (4.4%) | <.0001 |
| Cardiovascular diseases | 3449 (36.1%) | 2673 (11.1%) | <.0001 | 3895 (39.6%) | 3014 (12.9%) | <.0001 | 3499 (38.2%) | 2624 (12.5%) | <.0001 |
| Intellectual disability | 197 (2.1%) | 42 (0.2%) | <.0001 | 220 (2.2%) | 52 (0.2%) | <.0001 | 233 (2.5%) | 60 (0.3%) | <.0001 |
| Cancer | 1389 (14.5%) | 1295 (5.4%) | <.0001 | 1514 (15.4%) | 1396 (6.0%) | <.0001 | 1380 (15.1%) | 1407 (6.7%) | <.0001 |
| Time from first R568, G41 or G40 to first ASM (months) | 2.0±5.5 0.2 (-3.0 - 55.6) n=9561 | n=0 |  | 4.2±12.2 0.3 (-3.0 - 115.9) n=9843 | n=0 |  | 6.0±18.9 0.3 (-3.0 - 174.9) n=9165 | n=0 |  |
| ***Male - population*** | N=5311 | N=13864 |  | N=5692 | N=14121 |  | N=5545 | N=13329 |  |
| Age at inclusion | 58.8±20.4 63.0 (18.0 - 99.0) n=5311 | 57.9±20.5 62.0 (18.0 - 99.0) n=13864 | 0.0063 | 59.5±20.9 65.0 (18.0 - 99.0) n=5692 | 58.5±21.2 64.0 (18.0 - 99.0) n=14121 | 0.0028 | 59.7±21.5 66.0 (18.0 - 101.0) n=5545 | 58.6±21.8 65.0 (18.0 - 101.0) n=13329 | 0.0024 |
| Age at inclusion |  |  | 0.020 |  |  | 0.0079 |  |  | 0.0032 |
| <30 years | 727 (13.7%) | 2036 (14.7%) |  | 824 (14.5%) | 2229 (15.8%) |  | 894 (16.1%) | 2386 (17.9%) |  |
| 30-64 years | 2112 (39.8%) | 5606 (40.4%) |  | 1962 (34.5%) | 4928 (34.9%) |  | 1721 (31.0%) | 4138 (31.0%) |  |
| ≥65 years | 2472 (46.5%) | 6222 (44.9%) |  | 2906 (51.1%) | 6964 (49.3%) |  | 2930 (52.8%) | 6805 (51.1%) |  |
| Stroke | 1486 (28.0%) | 480 (3.5%) | <.0001 | 1599 (28.1%) | 577 (4.1%) | <.0001 | 1516 (27.3%) | 556 (4.2%) | <.0001 |
| TBI | 371 (7.0%) | 67 (0.5%) | <.0001 | 405 (7.1%) | 110 (0.8%) | <.0001 | 369 (6.7%) | 71 (0.5%) | <.0001 |
| Dementia | 230 (4.3%) | 125 (0.9%) | <.0001 | 268 (4.7%) | 140 (1.0%) | <.0001 | 237 (4.3%) | 125 (0.9%) | <.0001 |
| Brain tumour | 616 (11.6%) | 13 (0.1%) | <.0001 | 684 (12.0%) | 19 (0.1%) | <.0001 | 618 (11.1%) | 29 (0.2%) | <.0001 |
| Brain infection | 70 (1.3%) | 26 (0.2%) | <.0001 | 134 (2.4%) | 37 (0.3%) | <.0001 | 88 (1.6%) | 50 (0.4%) | <.0001 |
| Diabetes | 560 (10.5%) | 582 (4.2%) | <.0001 | 758 (13.3%) | 695 (4.9%) | <.0001 | 693 (12.5%) | 699 (5.2%) | <.0001 |
| Cardiovascular diseases | 2008 (37.8%) | 1738 (12.5%) | <.0001 | 2429 (42.7%) | 2086 (14.8%) | <.0001 | 2281 (41.1%) | 1896 (14.2%) | <.0001 |
| Intellectual disability | 123 (2.3%) | 28 (0.2%) | <.0001 | 126 (2.2%) | 38 (0.3%) | <.0001 | 143 (2.6%) | 41 (0.3%) | <.0001 |
| Cancer | 839 (15.8%) | 893 (6.4%) | <.0001 | 972 (17.1%) | 1030 (7.3%) | <.0001 | 903 (16.3%) | 1073 (8.1%) | <.0001 |
| Time from first R568, G41 or G40 to first ASM (months) | 2.1±5.7 0.2 (-3.0 - 54.3) n=5311 | n=0 |  | 4.5±12.8 0.3 (-3.0 - 113.3) n=5692 | n=0 |  | 6.5±19.4 0.3 (-3.0 - 174.9) n=5545 | n=0 |  |
| ***Female - population*** | N=4250 | N=10184 |  | N=4151 | N=9301 |  | N=3620 | N=7710 |  |
| Age at inclusion | 58.3±22.5 63.0 (18.0 - 101.0) n=4250 | 56.9±22.7 61.0 (18.0 - 101.0) n=10184 | 0.0011 | 58.3±23.2 64.0 (18.0 - 101.0) n=4151 | 56.5±23.4 61.0 (18.0 - 101.0) n=9301 | <.0001 | 58.1±23.4 64.0 (18.0 - 102.0) n=3620 | 56.4±23.8 62.0 (18.0 - 102.0) n=7710 | 0.0002 |
| Age at inclusion |  |  | 0.0043 |  |  | 0.0008 |  |  | 0.0008 |
| <30 years | 696 (16.4%) | 1844 (18.1%) |  | 739 (17.8%) | 1862 (20.0%) |  | 667 (18.4%) | 1647 (21.4%) |  |
| 30-64 years | 1570 (36.9%) | 3804 (37.4%) |  | 1395 (33.6%) | 3158 (34.0%) |  | 1158 (32.0%) | 2421 (31.4%) |  |
| ≥65 years | 1984 (46.7%) | 4536 (44.5%) |  | 2017 (48.6%) | 4281 (46.0%) |  | 1795 (49.6%) | 3642 (47.2%) |  |
| Stroke | 1060 (24.9%) | 300 (2.9%) | <.0001 | 1035 (24.9%) | 245 (2.6%) | <.0001 | 829 (22.9%) | 218 (2.8%) | <.0001 |
| TBI | 171 (4.0%) | 44 (0.4%) | <.0001 | 181 (4.4%) | 52 (0.6%) | <.0001 | 145 (4.0%) | 37 (0.5%) | <.0001 |
| Dementia | 229 (5.4%) | 134 (1.3%) | <.0001 | 243 (5.9%) | 113 (1.2%) | <.0001 | 178 (4.9%) | 90 (1.2%) | <.0001 |
| Brain tumour | 570 (13.4%) | 21 (0.2%) | <.0001 | 507 (12.2%) | 31 (0.3%) | <.0001 | 489 (13.5%) | 19 (0.2%) | <.0001 |
| Brain infection | 71 (1.7%) | 27 (0.3%) | <.0001 | 54 (1.3%) | 26 (0.3%) | <.0001 | 63 (1.7%) | 30 (0.4%) | <.0001 |
| Diabetes | 338 (8.0%) | 284 (2.8%) | <.0001 | 387 (9.3%) | 301 (3.2%) | <.0001 | 369 (10.2%) | 227 (2.9%) | <.0001 |
| Cardiovascular diseases | 1441 (33.9%) | 935 (9.2%) | <.0001 | 1466 (35.3%) | 928 (10.0%) | <.0001 | 1218 (33.6%) | 728 (9.4%) | <.0001 |
| Intellectual disability | 74 (1.7%) | 14 (0.1%) | <.0001 | 94 (2.3%) | 14 (0.2%) | <.0001 | 90 (2.5%) | 19 (0.2%) | <.0001 |
| Cancer | 550 (12.9%) | 402 (3.9%) | <.0001 | 542 (13.1%) | 366 (3.9%) | <.0001 | 477 (13.2%) | 334 (4.3%) | <.0001 |
| Time from first R568, G41 or G40 to first ASM (months) | 1.9±5.3 0.3 (-3.0 - 55.6) n=4250 | n=0 |  | 3.7±11.4 0.3 (-3.0 - 115.9) n=4151 | n=0 |  | 5.4±18.0 0.3 (-3.0 - 166.1) n=3620 | n=0 |  |
| Data are presented as mean±standard deviation, median (range) and number of observations, or number (percentage). For test between two groups Fisher’s exact test was used for binary variables, Mantel-Haenszel Chi-square trend test for ordered categorical variables, and Mann-Whitney U test for continuous variables. | | | | | | | | | |

### **eTable 2B. Baseline characteristics for cases and controls, overall and for calendar periods 2006-2010, 2011-2015, 2016-2020 overall and by sex (FAS population - Age at epilepsy onset <30 years)**

|  | **2006-2010 N=5303** | | | **2011-2015 N=5654** | | | **2016-2020 N=5594** | | |
| --- | --- | --- | --- | --- | --- | --- | --- | --- | --- |
| **Variable** | **Case N=1423** | **Control N=3880** | **p-value** | **Case N=1563** | **Control N=4091** | **p-value** | **Case N=1561** | **Control N=4033** | **p-value** |
| ***Overall - population*** |  |  |  |  |  |  |  |  |  |
| Age at inclusion | 22.8±3.4 22.0 (18.0 - 29.0) n=1423 | 22.8±3.4 22.0 (18.0 - 29.0) n=3880 | 0.92 | 22.8±3.4 23.0 (18.0 - 29.0) n=1563 | 22.8±3.4 22.0 (18.0 - 29.0) n=4091 | 0.47 | 23.0±3.4 23.0 (18.0 - 29.0) n=1561 | 22.9±3.4 23.0 (18.0 - 29.0) n=4033 | 0.32 |
| Sex |  |  | 0.39 |  |  | 0.24 |  |  | 0.20 |
| Male | 727 (51.1%) | 2036 (52.5%) |  | 824 (52.7%) | 2229 (54.5%) |  | 894 (57.3%) | 2386 (59.2%) |  |
| Female | 696 (48.9%) | 1844 (47.5%) |  | 739 (47.3%) | 1862 (45.5%) |  | 667 (42.7%) | 1647 (40.8%) |  |
| Stroke | 33 (2.3%) | 2 (0.1%) | <.0001 | 46 (2.9%) | 0 (0.0%) | <.0001 | 43 (2.8%) | 0 (0.0%) | <.0001 |
| TBI | 81 (5.7%) | 30 (0.8%) | <.0001 | 74 (4.7%) | 39 (1.0%) | <.0001 | 59 (3.8%) | 24 (0.6%) | <.0001 |
| Dementia | 2 (0.1%) | 0 (0.0%) | 0.07 | 1 (0.1%) | 0 (0.0%) | 0.28 | 1 (0.1%) | 0 (0.0%) | 0.28 |
| Brain tumour | 67 (4.7%) | 0 (0.0%) | <.0001 | 75 (4.8%) | 0 (0.0%) | <.0001 | 61 (3.9%) | 2 (0.0%) | <.0001 |
| Brain infection | 12 (0.8%) | 8 (0.2%) | 0.0018 | 21 (1.3%) | 10 (0.2%) | <.0001 | 22 (1.4%) | 17 (0.4%) | 0.0002 |
| Diabetes | 11 (0.8%) | 30 (0.8%) | 1.00 | 38 (2.4%) | 23 (0.6%) | <.0001 | 13 (0.8%) | 28 (0.7%) | 0.60 |
| Cardiovascular diseases | 83 (5.8%) | 31 (0.8%) | <.0001 | 83 (5.3%) | 34 (0.8%) | <.0001 | 57 (3.7%) | 24 (0.6%) | <.0001 |
| Intellectual disability | 111 (7.8%) | 22 (0.6%) | <.0001 | 133 (8.5%) | 21 (0.5%) | <.0001 | 153 (9.8%) | 41 (1.0%) | <.0001 |
| Cancer | 53 (3.7%) | 13 (0.3%) | <.0001 | 46 (2.9%) | 9 (0.2%) | <.0001 | 41 (2.6%) | 5 (0.1%) | <.0001 |
| Time from first R568, G41 or G40 to first ASM (months) | 3.4±7.4 0.5 (-2.9 - 55.6) n=1423 | n=0 |  | 5.5±12.1 0.9 (-3.0 - 106.3) n=1563 | n=0 |  | 6.4±13.8 1.2 (-3.0 - 119.4) n=1561 | n=0 |  |
| ***Male - population*** | N=727 | N=2036 |  | N=824 | N=2229 |  | N=894 | N=2386 |  |
| Age at inclusion | 22.8±3.5 23.0 (18.0 - 29.0) n=727 | 22.8±3.5 23.0 (18.0 - 29.0) n=2036 | 0.98 | 23.0±3.4 23.0 (18.0 - 29.0) n=824 | 22.9±3.4 23.0 (18.0 - 29.0) n=2229 | 0.78 | 23.2±3.4 23.0 (18.0 - 29.0) n=894 | 23.1±3.4 23.0 (18.0 - 29.0) n=2386 | 0.58 |
| Stroke | 17 (2.3%) | 1 (0.0%) | <.0001 | 30 (3.6%) | 0 (0.0%) | <.0001 | 33 (3.7%) | 0 (0.0%) | <.0001 |
| TBI | 54 (7.4%) | 20 (1.0%) | <.0001 | 53 (6.4%) | 25 (1.1%) | <.0001 | 42 (4.7%) | 18 (0.8%) | <.0001 |
| Dementia | 2 (0.3%) | 0 (0.0%) | 0.07 | 1 (0.1%) | 0 (0.0%) | 0.27 | 0 (0%) | 0 (0%) |  |
| Brain tumour | 36 (5.0%) | 0 (0.0%) | <.0001 | 48 (5.8%) | 0 (0.0%) | <.0001 | 43 (4.8%) | 2 (0.1%) | <.0001 |
| Brain infection | 6 (0.8%) | 4 (0.2%) | 0.025 | 17 (2.1%) | 8 (0.4%) | <.0001 | 15 (1.7%) | 10 (0.4%) | 0.0009 |
| Diabetes | 5 (0.7%) | 20 (1.0%) | 0.65 | 23 (2.8%) | 12 (0.5%) | <.0001 | 11 (1.2%) | 14 (0.6%) | 0.07 |
| Cardiovascular diseases | 43 (5.9%) | 20 (1.0%) | <.0001 | 51 (6.2%) | 20 (0.9%) | <.0001 | 31 (3.5%) | 17 (0.7%) | <.0001 |
| Intellectual disability | 70 (9.6%) | 13 (0.6%) | <.0001 | 81 (9.8%) | 16 (0.7%) | <.0001 | 96 (10.7%) | 27 (1.1%) | <.0001 |
| Cancer | 28 (3.9%) | 7 (0.3%) | <.0001 | 30 (3.6%) | 7 (0.3%) | <.0001 | 28 (3.1%) | 4 (0.2%) | <.0001 |
| Time from first R568, G41 or G40 to first ASM (months) | 3.5±7.7 0.6 (-2.9 - 50.4) n=727 | n=0 |  | 6.4±12.8 1.3 (-3.0 - 102.9) n=824 | n=0 |  | 7.1±14.9 1.5 (-3.0 - 119.4) n=894 | n=0 |  |
| ***Female - population*** | N=696 | N=1844 |  | N=739 | N=1862 |  | N=667 | N=1647 |  |
| Age at inclusion | 22.8±3.4 22.0 (18.0 - 29.0) n=696 | 22.7±3.3 22.0 (18.0 - 29.0) n=1844 | 0.90 | 22.7±3.4 22.0 (18.0 - 29.0) n=739 | 22.6±3.4 22.0 (18.0 - 29.0) n=1862 | 0.40 | 22.8±3.4 23.0 (18.0 - 29.0) n=667 | 22.7±3.4 23.0 (18.0 - 29.0) n=1647 | 0.32 |
| Stroke | 16 (2.3%) | 1 (0.1%) | <.0001 | 16 (2.2%) | 0 (0.0%) | <.0001 | 10 (1.5%) | 0 (0.0%) | <.0001 |
| TBI | 27 (3.9%) | 10 (0.5%) | <.0001 | 21 (2.8%) | 14 (0.8%) | <.0001 | 17 (2.5%) | 6 (0.4%) | <.0001 |
| Dementia | 0 (0%) | 0 (0%) |  | 0 (0%) | 0 (0%) |  | 1 (0.1%) | 0 (0.0%) | 0.29 |
| Brain tumour | 31 (4.5%) | 0 (0.0%) | <.0001 | 27 (3.7%) | 0 (0.0%) | <.0001 | 18 (2.7%) | 0 (0.0%) | <.0001 |
| Brain infection | 6 (0.9%) | 4 (0.2%) | 0.031 | 4 (0.5%) | 2 (0.1%) | 0.06 | 7 (1.0%) | 7 (0.4%) | 0.13 |
| Diabetes | 6 (0.9%) | 10 (0.5%) | 0.40 | 15 (2.0%) | 11 (0.6%) | 0.0018 | 2 (0.3%) | 14 (0.9%) | 0.18 |
| Cardiovascular diseases | 40 (5.7%) | 11 (0.6%) | <.0001 | 32 (4.3%) | 14 (0.8%) | <.0001 | 26 (3.9%) | 7 (0.4%) | <.0001 |
| Intellectual disability | 41 (5.9%) | 9 (0.5%) | <.0001 | 52 (7.0%) | 5 (0.3%) | <.0001 | 57 (8.5%) | 14 (0.9%) | <.0001 |
| Cancer | 25 (3.6%) | 6 (0.3%) | <.0001 | 16 (2.2%) | 2 (0.1%) | <.0001 | 13 (1.9%) | 1 (0.1%) | <.0001 |
| Time from first R568, G41 or G40 to first ASM (months) | 3.2±7.1 0.4 (-2.9 - 55.6) n=696 | n=0 |  | 4.5±11.1 0.7 (-2.8 - 106.3) n=739 | n=0 |  | 5.5±12.3 0.9 (-2.9 - 104.8) n=667 | n=0 |  |
| Data are presented as mean±standard deviation, median (range) and number of observations, or number (percentage). For test between two groups Fisher’s exact test was used for binary variables and Mann-Whitney U test for continuous variables. | | | | | | | | | |

### **eTable 2C. Baseline characteristics for cases and controls, overall and for calendar periods 2006-2010, 2011-2015, 2016-2020 overall and by sex (FAS population - Age at epilepsy onset 30-64 years)**

|  | **2006-2010 N=13092** | | | **2011-2015 N=11443** | | | **2016-2020 N=9438** | | |
| --- | --- | --- | --- | --- | --- | --- | --- | --- | --- |
| **Variable** | **Case N=3682** | **Control N=9410** | **p-value** | **Case N=3357** | **Control N=8086** | **p-value** | **Case N=2879** | **Control N=6559** | **p-value** |
| ***Overall - population*** |  |  |  |  |  |  |  |  |  |
| Age at inclusion | 49.6±10.4 51.0 (30.0 - 64.0) n=3682 | 49.5±10.4 51.0 (30.0 - 64.0) n=9410 | 0.48 | 48.9±10.5 50.0 (30.0 - 64.0) n=3357 | 48.7±10.6 50.0 (30.0 - 64.0) n=8086 | 0.37 | 48.7±10.7 50.0 (30.0 - 64.0) n=2879 | 48.6±10.7 50.0 (30.0 - 64.0) n=6559 | 0.51 |
| Sex |  |  | 0.021 |  |  | 0.013 |  |  | 0.0024 |
| Male | 2112 (57.4%) | 5606 (59.6%) |  | 1962 (58.4%) | 4928 (60.9%) |  | 1721 (59.8%) | 4138 (63.1%) |  |
| Female | 1570 (42.6%) | 3804 (40.4%) |  | 1395 (41.6%) | 3158 (39.1%) |  | 1158 (40.2%) | 2421 (36.9%) |  |
| Stroke | 635 (17.2%) | 68 (0.7%) | <.0001 | 519 (15.5%) | 46 (0.6%) | <.0001 | 427 (14.8%) | 48 (0.7%) | <.0001 |
| TBI | 227 (6.2%) | 19 (0.2%) | <.0001 | 203 (6.0%) | 30 (0.4%) | <.0001 | 156 (5.4%) | 20 (0.3%) | <.0001 |
| Dementia | 47 (1.3%) | 3 (0.0%) | <.0001 | 49 (1.5%) | 2 (0.0%) | <.0001 | 32 (1.1%) | 3 (0.0%) | <.0001 |
| Brain tumour | 648 (17.6%) | 8 (0.1%) | <.0001 | 595 (17.7%) | 12 (0.1%) | <.0001 | 481 (16.7%) | 8 (0.1%) | <.0001 |
| Brain infection | 84 (2.3%) | 23 (0.2%) | <.0001 | 91 (2.7%) | 34 (0.4%) | <.0001 | 62 (2.2%) | 28 (0.4%) | <.0001 |
| Diabetes | 226 (6.1%) | 158 (1.7%) | <.0001 | 223 (6.6%) | 155 (1.9%) | <.0001 | 193 (6.7%) | 124 (1.9%) | <.0001 |
| Cardiovascular diseases | 808 (21.9%) | 395 (4.2%) | <.0001 | 739 (22.0%) | 345 (4.3%) | <.0001 | 605 (21.0%) | 274 (4.2%) | <.0001 |
| Intellectual disability | 69 (1.9%) | 11 (0.1%) | <.0001 | 67 (2.0%) | 21 (0.3%) | <.0001 | 61 (2.1%) | 10 (0.2%) | <.0001 |
| Cancer | 544 (14.8%) | 201 (2.1%) | <.0001 | 486 (14.5%) | 165 (2.0%) | <.0001 | 388 (13.5%) | 131 (2.0%) | <.0001 |
| Time from first R568, G41 or G40 to first ASM (months) | 2.2±5.6 0.2 (-3.0 - 50.0) n=3682 | n=0 |  | 5.5±14.6 0.3 (-3.0 - 115.9) n=3357 | n=0 |  | 8.4±23.2 0.3 (-3.0 - 174.9) n=2879 | n=0 |  |
| ***Male - population*** | N=2112 | N=5606 |  | N=1962 | N=4928 |  | N=1721 | N=4138 |  |
| Age at inclusion | 50.5±10.0 52.0 (30.0 - 64.0) n=2112 | 50.3±10.1 52.0 (30.0 - 64.0) n=5606 | 0.61 | 49.7±10.4 52.0 (30.0 - 64.0) n=1962 | 49.5±10.5 51.0 (30.0 - 64.0) n=4928 | 0.59 | 49.9±10.5 52.0 (30.0 - 64.0) n=1721 | 49.8±10.6 52.0 (30.0 - 64.0) n=4138 | 0.54 |
| Stroke | 428 (20.3%) | 48 (0.9%) | <.0001 | 335 (17.1%) | 36 (0.7%) | <.0001 | 313 (18.2%) | 39 (0.9%) | <.0001 |
| TBI | 164 (7.8%) | 8 (0.1%) | <.0001 | 143 (7.3%) | 19 (0.4%) | <.0001 | 119 (6.9%) | 15 (0.4%) | <.0001 |
| Dementia | 22 (1.0%) | 3 (0.1%) | <.0001 | 24 (1.2%) | 2 (0.0%) | <.0001 | 19 (1.1%) | 2 (0.0%) | <.0001 |
| Brain tumour | 330 (15.6%) | 3 (0.1%) | <.0001 | 344 (17.5%) | 6 (0.1%) | <.0001 | 285 (16.6%) | 6 (0.1%) | <.0001 |
| Brain infection | 37 (1.8%) | 11 (0.2%) | <.0001 | 70 (3.6%) | 20 (0.4%) | <.0001 | 33 (1.9%) | 20 (0.5%) | <.0001 |
| Diabetes | 161 (7.6%) | 115 (2.1%) | <.0001 | 154 (7.8%) | 114 (2.3%) | <.0001 | 126 (7.3%) | 98 (2.4%) | <.0001 |
| Cardiovascular diseases | 536 (25.4%) | 274 (4.9%) | <.0001 | 495 (25.2%) | 254 (5.2%) | <.0001 | 424 (24.6%) | 200 (4.8%) | <.0001 |
| Intellectual disability | 42 (2.0%) | 6 (0.1%) | <.0001 | 31 (1.6%) | 15 (0.3%) | <.0001 | 34 (2.0%) | 5 (0.1%) | <.0001 |
| Cancer | 303 (14.3%) | 123 (2.2%) | <.0001 | 293 (14.9%) | 100 (2.0%) | <.0001 | 242 (14.1%) | 84 (2.0%) | <.0001 |
| Time from first R568, G41 or G40 to first ASM (months) | 2.3±5.5 0.2 (-3.0 - 50.0) n=2112 | n=0 |  | 5.6±14.8 0.3 (-3.0 - 105.9) n=1962 | n=0 |  | 8.6±23.5 0.3 (-3.0 - 174.9) n=1721 | n=0 |  |
| ***Female - population*** | N=1570 | N=3804 |  | N=1395 | N=3158 |  | N=1158 | N=2421 |  |
| Age at inclusion | 48.4±10.7 49.0 (30.0 - 64.0) n=1570 | 48.2±10.8 49.0 (30.0 - 64.0) n=3804 | 0.41 | 47.7±10.6 48.0 (30.0 - 64.0) n=1395 | 47.3±10.7 48.0 (30.0 - 64.0) n=3158 | 0.26 | 46.9±10.6 47.0 (30.0 - 64.0) n=1158 | 46.5±10.6 46.0 (30.0 - 64.0) n=2421 | 0.30 |
| Stroke | 207 (13.2%) | 20 (0.5%) | <.0001 | 184 (13.2%) | 10 (0.3%) | <.0001 | 114 (9.8%) | 9 (0.4%) | <.0001 |
| TBI | 63 (4.0%) | 11 (0.3%) | <.0001 | 60 (4.3%) | 11 (0.3%) | <.0001 | 37 (3.2%) | 5 (0.2%) | <.0001 |
| Dementia | 25 (1.6%) | 0 (0.0%) | <.0001 | 25 (1.8%) | 0 (0.0%) | <.0001 | 13 (1.1%) | 1 (0.0%) | <.0001 |
| Brain tumour | 318 (20.3%) | 5 (0.1%) | <.0001 | 251 (18.0%) | 6 (0.2%) | <.0001 | 196 (16.9%) | 2 (0.1%) | <.0001 |
| Brain infection | 47 (3.0%) | 12 (0.3%) | <.0001 | 21 (1.5%) | 14 (0.4%) | 0.0003 | 29 (2.5%) | 8 (0.3%) | <.0001 |
| Diabetes | 65 (4.1%) | 43 (1.1%) | <.0001 | 69 (4.9%) | 41 (1.3%) | <.0001 | 67 (5.8%) | 26 (1.1%) | <.0001 |
| Cardiovascular diseases | 272 (17.3%) | 121 (3.2%) | <.0001 | 244 (17.5%) | 91 (2.9%) | <.0001 | 181 (15.6%) | 74 (3.1%) | <.0001 |
| Intellectual disability | 27 (1.7%) | 5 (0.1%) | <.0001 | 36 (2.6%) | 6 (0.2%) | <.0001 | 27 (2.3%) | 5 (0.2%) | <.0001 |
| Cancer | 241 (15.4%) | 78 (2.1%) | <.0001 | 193 (13.8%) | 65 (2.1%) | <.0001 | 146 (12.6%) | 47 (1.9%) | <.0001 |
| Time from first R568, G41 or G40 to first ASM (months) | 2.1±5.6 0.2 (-3.0 - 49.8) n=1570 | n=0 |  | 5.2±14.5 0.3 (-3.0 - 115.9) n=1395 | n=0 |  | 8.2±22.8 0.3 (-2.9 - 166.1) n=1158 | n=0 |  |
| Data are presented as mean±standard deviation, median (range) and number of observations, or number (percentage). For test between two groups Fisher’s exact test was used for binary variables and Mann-Whitney U test for continuous variables. | | | | | | | | | |

### **eTable 2D. Baseline characteristics for cases and controls, overall and for calendar periods 2006-2010, 2011-2015, 2016-2020 overall and by sex (FAS population - Age at epilepsy onset ≥65 years)**

|  | **2006-2010 N=15214** | | | **2011-2015 N=16168** | | | **2016-2020 N=15172** | | |
| --- | --- | --- | --- | --- | --- | --- | --- | --- | --- |
| **Variable** | **Case N=4456** | **Control N=10758** | **p-value** | **Case N=4923** | **Control N=11245** | **p-value** | **Case N=4725** | **Control N=10447** | **p-value** |
| ***Overall - population*** |  |  |  |  |  |  |  |  |  |
| Age at inclusion | 77.4±7.7 77.0 (65.0 - 101.0) n=4456 | 77.0±7.6 77.0 (65.0 - 101.0) n=10758 | 0.0100 | 77.3±7.8 77.0 (65.0 - 101.0) n=4923 | 76.9±7.7 76.0 (65.0 - 101.0) n=11245 | 0.0022 | 77.4±7.6 77.0 (65.0 - 102.0) n=4725 | 77.1±7.5 76.0 (65.0 - 102.0) n=10447 | 0.031 |
| Sex |  |  | 0.0077 |  |  | 0.0005 |  |  | 0.0002 |
| Male | 2472 (55.5%) | 6222 (57.8%) |  | 2906 (59.0%) | 6964 (61.9%) |  | 2930 (62.0%) | 6805 (65.1%) |  |
| Female | 1984 (44.5%) | 4536 (42.2%) |  | 2017 (41.0%) | 4281 (38.1%) |  | 1795 (38.0%) | 3642 (34.9%) |  |
| Stroke | 1878 (42.1%) | 710 (6.6%) | <.0001 | 2069 (42.0%) | 776 (6.9%) | <.0001 | 1875 (39.7%) | 726 (6.9%) | <.0001 |
| TBI | 234 (5.3%) | 62 (0.6%) | <.0001 | 309 (6.3%) | 93 (0.8%) | <.0001 | 299 (6.3%) | 64 (0.6%) | <.0001 |
| Dementia | 410 (9.2%) | 256 (2.4%) | <.0001 | 461 (9.4%) | 251 (2.2%) | <.0001 | 382 (8.1%) | 212 (2.0%) | <.0001 |
| Brain tumour | 471 (10.6%) | 26 (0.2%) | <.0001 | 521 (10.6%) | 38 (0.3%) | <.0001 | 565 (12.0%) | 38 (0.4%) | <.0001 |
| Brain infection | 45 (1.0%) | 22 (0.2%) | <.0001 | 76 (1.5%) | 19 (0.2%) | <.0001 | 67 (1.4%) | 35 (0.3%) | <.0001 |
| Diabetes | 661 (14.8%) | 678 (6.3%) | <.0001 | 884 (18.0%) | 818 (7.3%) | <.0001 | 856 (18.1%) | 774 (7.4%) | <.0001 |
| Cardiovascular diseases | 2558 (57.4%) | 2247 (20.9%) | <.0001 | 3073 (62.4%) | 2635 (23.4%) | <.0001 | 2837 (60.0%) | 2326 (22.3%) | <.0001 |
| Intellectual disability | 17 (0.4%) | 9 (0.1%) | 0.0001 | 20 (0.4%) | 10 (0.1%) | <.0001 | 19 (0.4%) | 9 (0.1%) | <.0001 |
| Cancer | 792 (17.8%) | 1081 (10.0%) | <.0001 | 982 (19.9%) | 1222 (10.9%) | <.0001 | 951 (20.1%) | 1271 (12.2%) | <.0001 |
| Time from first R568, G41 or G40 to first ASM (months) | 1.4±4.6 0.3 (-3.0 - 54.4) n=4456 | n=0 |  | 2.9±10.1 0.3 (-3.0 - 113.3) n=4923 | n=0 |  | 4.5±17.1 0.2 (-3.0 - 172.4) n=4725 | n=0 |  |
| ***Male - population*** | N=2472 | N=6222 |  | N=2906 | N=6964 |  | N=2930 | N=6805 |  |
| Age at inclusion | 76.5±7.4 76.0 (65.0 - 99.0) n=2472 | 76.2±7.3 76.0 (65.0 - 99.0) n=6222 | 0.11 | 76.4±7.4 76.0 (65.0 - 99.0) n=2906 | 76.1±7.3 75.0 (65.0 - 99.0) n=6964 | 0.15 | 76.7±7.2 76.0 (65.0 - 101.0) n=2930 | 76.5±7.0 76.0 (65.0 - 101.0) n=6805 | 0.34 |
| Stroke | 1041 (42.1%) | 431 (6.9%) | <.0001 | 1234 (42.5%) | 541 (7.8%) | <.0001 | 1170 (39.9%) | 517 (7.6%) | <.0001 |
| TBI | 153 (6.2%) | 39 (0.6%) | <.0001 | 209 (7.2%) | 66 (0.9%) | <.0001 | 208 (7.1%) | 38 (0.6%) | <.0001 |
| Dementia | 206 (8.3%) | 122 (2.0%) | <.0001 | 243 (8.4%) | 138 (2.0%) | <.0001 | 218 (7.4%) | 123 (1.8%) | <.0001 |
| Brain tumour | 250 (10.1%) | 10 (0.2%) | <.0001 | 292 (10.0%) | 13 (0.2%) | <.0001 | 290 (9.9%) | 21 (0.3%) | <.0001 |
| Brain infection | 27 (1.1%) | 11 (0.2%) | <.0001 | 47 (1.6%) | 9 (0.1%) | <.0001 | 40 (1.4%) | 20 (0.3%) | <.0001 |
| Diabetes | 394 (15.9%) | 447 (7.2%) | <.0001 | 581 (20.0%) | 569 (8.2%) | <.0001 | 556 (19.0%) | 587 (8.6%) | <.0001 |
| Cardiovascular diseases | 1429 (57.8%) | 1444 (23.2%) | <.0001 | 1883 (64.8%) | 1812 (26.0%) | <.0001 | 1826 (62.3%) | 1679 (24.7%) | <.0001 |
| Intellectual disability | 11 (0.4%) | 9 (0.1%) | 0.012 | 14 (0.5%) | 7 (0.1%) | 0.0005 | 13 (0.4%) | 9 (0.1%) | 0.0048 |
| Cancer | 508 (20.6%) | 763 (12.3%) | <.0001 | 649 (22.3%) | 923 (13.3%) | <.0001 | 633 (21.6%) | 985 (14.5%) | <.0001 |
| Time from first R568, G41 or G40 to first ASM (months) | 1.6±5.0 0.2 (-3.0 - 54.3) n=2472 | n=0 |  | 3.2±11.1 0.3 (-3.0 - 113.3) n=2906 | n=0 |  | 5.0±17.7 0.2 (-3.0 - 172.4) n=2930 | n=0 |  |
| ***Female - population*** | N=1984 | N=4536 |  | N=2017 | N=4281 |  | N=1795 | N=3642 |  |
| Age at inclusion | 78.5±7.9 78.0 (65.0 - 101.0) n=1984 | 78.2±7.9 78.0 (65.0 - 101.0) n=4536 | 0.11 | 78.7±8.3 78.0 (65.0 - 101.0) n=2017 | 78.1±8.2 78.0 (65.0 - 101.0) n=4281 | 0.017 | 78.5±8.2 78.0 (65.0 - 102.0) n=1795 | 78.1±8.1 77.0 (65.0 - 102.0) n=3642 | 0.09 |
| Stroke | 837 (42.2%) | 279 (6.2%) | <.0001 | 835 (41.4%) | 235 (5.5%) | <.0001 | 705 (39.3%) | 209 (5.7%) | <.0001 |
| TBI | 81 (4.1%) | 23 (0.5%) | <.0001 | 100 (5.0%) | 27 (0.6%) | <.0001 | 91 (5.1%) | 26 (0.7%) | <.0001 |
| Dementia | 204 (10.3%) | 134 (3.0%) | <.0001 | 218 (10.8%) | 113 (2.6%) | <.0001 | 164 (9.1%) | 89 (2.4%) | <.0001 |
| Brain tumour | 221 (11.1%) | 16 (0.4%) | <.0001 | 229 (11.4%) | 25 (0.6%) | <.0001 | 275 (15.3%) | 17 (0.5%) | <.0001 |
| Brain infection | 18 (0.9%) | 11 (0.2%) | 0.0004 | 29 (1.4%) | 10 (0.2%) | <.0001 | 27 (1.5%) | 15 (0.4%) | <.0001 |
| Diabetes | 267 (13.5%) | 231 (5.1%) | <.0001 | 303 (15.0%) | 249 (5.8%) | <.0001 | 300 (16.7%) | 187 (5.1%) | <.0001 |
| Cardiovascular diseases | 1129 (56.9%) | 803 (17.7%) | <.0001 | 1190 (59.0%) | 823 (19.2%) | <.0001 | 1011 (56.3%) | 647 (17.8%) | <.0001 |
| Intellectual disability | 6 (0.3%) | 0 (0.0%) | 0.0008 | 6 (0.3%) | 3 (0.1%) | 0.035 | 6 (0.3%) | 0 (0.0%) | 0.0013 |
| Cancer | 284 (14.3%) | 318 (7.0%) | <.0001 | 333 (16.5%) | 299 (7.0%) | <.0001 | 318 (17.7%) | 286 (7.9%) | <.0001 |
| Time from first R568, G41 or G40 to first ASM (months) | 1.2±4.1 0.3 (-3.0 - 54.4) n=1984 | n=0 |  | 2.4±8.5 0.3 (-3.0 - 96.2) n=2017 | n=0 |  | 3.6±16.0 0.2 (-3.0 - 164.7) n=1795 | n=0 |  |
| Data are presented as mean±standard deviation, median (range) and number of observations, or number (percentage). For test between two groups Fisher’s exact test was used for binary variables and Mann-Whitney U test for continuous variables. | | | | | | | | | |

### **eTable 3A. Unadjusted, and fully adjusted HR, CIs, p-values for new-onset depression from Cox regression in cases vs controls during periods 2006-2010, 2011-2015 and 2016-2020 overall and by sex (FAS population)**

|  | | | **Case** | | | **Control** | | | **Case vs. Control** | | **Interaction** | |
| --- | --- | --- | --- | --- | --- | --- | --- | --- | --- | --- | --- | --- |
| **Subgroup** | **Period** | **Period** | **n/N (%) events** | **Follow-up time Median (IQR) Sum** | **Event rate (95% CI) per 100 person- years** | **n/N (%) events** | **Follow-up time Median (IQR) Sum** | **Event rate (95% CI) per 100 person- years** | **Hazard ratio (95% CI) p-value** | *** Hazard ratio (95% CI) p-value** | **p-value** | *** p-value** |
| ***Overall - population*** |  |  |  |  |  |  |  |  |  |  |  |  |
|  | **2006-2010 vs. 2016-2020** | **2006-2010** | 1986/9561 (20.8%) | 3.75 (1.34-5.48) Sum=34398 | 5.8 (5.5-6.0) | 2648/24048 (11.0%) | 4.87 (3.60-6.25) Sum=115464 | 2.3 (2.2-2.4) | 2.45 (2.31 - 2.59) p=<.0001 | 1.92 (1.79 - 2.06) p=<.0001 |  |  |
|  |  | **2016-2020** | 2020/9165 (22.0%) | 3.90 (1.57-5.72) Sum=34627 | 5.8 (5.6-6.1) | 2560/21039 (12.2%) | 4.94 (3.59-6.41) Sum=102371 | 2.5 (2.4-2.6) | 2.28 (2.16 - 2.42) p=<.0001 | 1.81 (1.69 - 1.94) p=<.0001 | 0.08 | 0.06 |
|  | **2006-2010 vs. 2011-2015** | **2006-2010** | 1986/9561 (20.8%) | 3.75 (1.34-5.48) Sum=34398 | 5.8 (5.5-6.0) | 2648/24048 (11.0%) | 4.87 (3.60-6.25) Sum=115464 | 2.3 (2.2-2.4) | 2.45 (2.31 - 2.59) p=<.0001 | 1.92 (1.79 - 2.06) p=<.0001 |  |  |
|  |  | **2011-2015** | 2073/9843 (21.1%) | 3.81 (1.45-5.62) Sum=36394 | 5.7 (5.5-5.9) | 2701/23422 (11.5%) | 4.93 (3.60-6.38) Sum=113671 | 2.4 (2.3-2.5) | 2.33 (2.20 - 2.47) p=<.0001 | 1.84 (1.72 - 1.97) p=<.0001 | 0.25 | 0.12 |
|  | **2011-2015 vs. 2016-2020** | **2011-2015** | 2073/9843 (21.1%) | 3.81 (1.45-5.62) Sum=36394 | 5.7 (5.5-5.9) | 2701/23422 (11.5%) | 4.93 (3.60-6.38) Sum=113671 | 2.4 (2.3-2.5) | 2.33 (2.20 - 2.47) p=<.0001 | 1.84 (1.72 - 1.97) p=<.0001 |  |  |
|  |  | **2016-2020** | 2020/9165 (22.0%) | 3.90 (1.57-5.72) Sum=34627 | 5.8 (5.6-6.1) | 2560/21039 (12.2%) | 4.94 (3.59-6.41) Sum=102371 | 2.5 (2.4-2.6) | 2.28 (2.16 - 2.42) p=<.0001 | 1.81 (1.69 - 1.94) p=<.0001 | 0.55 | 0.71 |
| ***Male - population*** |  |  |  |  |  |  |  |  |  |  |  |  |
|  | **2006-2010 vs. 2016-2020** | **2006-2010** | 1019/5311 (19.2%) | 3.78 (1.39-5.51) Sum=19277 | 5.3 (5.0-5.6) | 1248/13864 (9.0%) | 4.93 (3.65-6.28) Sum=67326 | 1.9 (1.8-2.0) | 2.76 (2.54 - 3.00) p=<.0001 | 2.11 (1.91 - 2.33) p=<.0001 |  |  |
|  |  | **2016-2020** | 1129/5545 (20.4%) | 3.94 (1.65-5.75) Sum=21122 | 5.3 (5.0-5.7) | 1395/13329 (10.5%) | 4.95 (3.62-6.38) Sum=65047 | 2.1 (2.0-2.3) | 2.44 (2.26 - 2.64) p=<.0001 | 1.89 (1.73 - 2.08) p=<.0001 | 0.025 | 0.018 |
|  | **2006-2010 vs. 2011-2015** | **2006-2010** | 1019/5311 (19.2%) | 3.78 (1.39-5.51) Sum=19277 | 5.3 (5.0-5.6) | 1248/13864 (9.0%) | 4.93 (3.65-6.28) Sum=67326 | 1.9 (1.8-2.0) | 2.76 (2.54 - 3.00) p=<.0001 | 2.11 (1.91 - 2.33) p=<.0001 |  |  |
|  |  | **2011-2015** | 1107/5692 (19.4%) | 3.87 (1.48-5.66) Sum=21233 | 5.2 (4.9-5.5) | 1354/14121 (9.6%) | 4.96 (3.64-6.41) Sum=68938 | 2.0 (1.9-2.1) | 2.58 (2.38 - 2.79) p=<.0001 | 1.98 (1.80 - 2.18) p=<.0001 | 0.24 | 0.09 |
|  | **2011-2015 vs. 2016-2020** | **2011-2015** | 1107/5692 (19.4%) | 3.87 (1.48-5.66) Sum=21233 | 5.2 (4.9-5.5) | 1354/14121 (9.6%) | 4.96 (3.64-6.41) Sum=68938 | 2.0 (1.9-2.1) | 2.58 (2.38 - 2.79) p=<.0001 | 1.98 (1.80 - 2.18) p=<.0001 |  |  |
|  |  | **2016-2020** | 1129/5545 (20.4%) | 3.94 (1.65-5.75) Sum=21122 | 5.3 (5.0-5.7) | 1395/13329 (10.5%) | 4.95 (3.62-6.38) Sum=65047 | 2.1 (2.0-2.3) | 2.44 (2.26 - 2.64) p=<.0001 | 1.89 (1.73 - 2.08) p=<.0001 | 0.29 | 0.50 |
| ***Female - population*** |  |  |  |  |  |  |  |  |  |  |  |  |
|  | **2006-2010 vs. 2016-2020** | **2006-2010** | 967/4250 (22.8%) | 3.70 (1.26-5.44) Sum=15120 | 6.4 (6.0-6.8) | 1400/10184 (13.7%) | 4.79 (3.53-6.22) Sum=48138 | 2.9 (2.8-3.1) | 2.14 (1.97 - 2.33) p=<.0001 | 1.77 (1.61 - 1.95) p=<.0001 |  |  |
|  |  | **2016-2020** | 891/3620 (24.6%) | 3.83 (1.44-5.66) Sum=13505 | 6.6 (6.2-7.0) | 1165/7710 (15.1%) | 4.91 (3.54-6.43) Sum=37324 | 3.1 (2.9-3.3) | 2.07 (1.90 - 2.26) p=<.0001 | 1.73 (1.56 - 1.91) p=<.0001 | 0.55 | 0.52 |
|  | **2006-2010 vs. 2011-2015** | **2006-2010** | 967/4250 (22.8%) | 3.70 (1.26-5.44) Sum=15120 | 6.4 (6.0-6.8) | 1400/10184 (13.7%) | 4.79 (3.53-6.22) Sum=48138 | 2.9 (2.8-3.1) | 2.14 (1.97 - 2.33) p=<.0001 | 1.77 (1.61 - 1.95) p=<.0001 |  |  |
|  |  | **2011-2015** | 966/4151 (23.3%) | 3.73 (1.40-5.55) Sum=15162 | 6.4 (6.0-6.8) | 1347/9301 (14.5%) | 4.90 (3.54-6.36) Sum=44733 | 3.0 (2.9-3.2) | 2.07 (1.90 - 2.25) p=<.0001 | 1.71 (1.55 - 1.88) p=<.0001 | 0.52 | 0.45 |
|  | **2011-2015 vs. 2016-2020** | **2011-2015** | 966/4151 (23.3%) | 3.73 (1.40-5.55) Sum=15162 | 6.4 (6.0-6.8) | 1347/9301 (14.5%) | 4.90 (3.54-6.36) Sum=44733 | 3.0 (2.9-3.2) | 2.07 (1.90 - 2.25) p=<.0001 | 1.71 (1.55 - 1.88) p=<.0001 |  |  |
|  |  | **2016-2020** | 891/3620 (24.6%) | 3.83 (1.44-5.66) Sum=13505 | 6.6 (6.2-7.0) | 1165/7710 (15.1%) | 4.91 (3.54-6.43) Sum=37324 | 3.1 (2.9-3.3) | 2.07 (1.90 - 2.26) p=<.0001 | 1.73 (1.56 - 1.91) p=<.0001 | 0.99 | 0.95 |
| Confidence interval for unadjusted event rates per 100 person years are obtained from exact Poisson confidence limits. Cox regression was used for time to any event presented by HR. * Adjusted for Age at inclusion, Sex, Stroke, TBI, Dementia, Brain tumour, Brain infection, Diabetes, Cardiovascular diseases, Intellectual disability, Cancer. The p-value for interaction was calculated between study group and each pairwise time period. | | | | | | | | | | | | |

### **eTable 3B. Unadjusted, and fully adjusted HR, CIs, p-values for new-onset depression from Cox regression in cases vs controls during periods 2006-2010, 2011-2015 and 2016-2020 for each subgroup overall and by sex (FAS population)**

|  | | | **Case** | | | **Control** | | | **Case vs. Control** | | **Interaction** | |
| --- | --- | --- | --- | --- | --- | --- | --- | --- | --- | --- | --- | --- |
| **Subgroup** | **Period** | **Period** | **n/N (%) events** | **Follow-up time Median (IQR) Sum** | **Event rate (95% CI) per 100 person- years** | **n/N (%) events** | **Follow-up time Median (IQR) Sum** | **Event rate (95% CI) per 100 person- years** | **Hazard ratio (95% CI) p-value** | *** Hazard ratio (95% CI) p-value** | **p-value** | *** p-value** |
| ***Overall - population*** |  |  |  |  |  |  |  |  |  |  |  |  |
| **<30 years of age at inclusion** | **2006-2010 vs. 2016-2020** | **2006-2010** | 259/1423 (18.2%) | 4.65 (3.49-6.05) Sum=6552 | 4.0 (3.5-4.5) | 314/3880 (8.1%) | 5.01 (3.89-6.35) Sum=19731 | 1.6 (1.4-1.8) | 2.48 (2.11 - 2.93) p=<.0001 | 2.33 (1.96 - 2.78) p=<.0001 |  |  |
|  |  | **2016-2020** | 284/1561 (18.2%) | 4.92 (3.50-6.47) Sum=7469 | 3.8 (3.4-4.3) | 447/4033 (11.1%) | 5.25 (3.92-6.61) Sum=20789 | 2.2 (2.0-2.4) | 1.76 (1.52 - 2.04) p=<.0001 | 1.63 (1.39 - 1.91) p=<.0001 | 0.0028 | 0.0036 |
|  | **2006-2010 vs. 2011-2015** | **2006-2010** | 259/1423 (18.2%) | 4.65 (3.49-6.05) Sum=6552 | 4.0 (3.5-4.5) | 314/3880 (8.1%) | 5.01 (3.89-6.35) Sum=19731 | 1.6 (1.4-1.8) | 2.48 (2.11 - 2.93) p=<.0001 | 2.33 (1.96 - 2.78) p=<.0001 |  |  |
|  |  | **2011-2015** | 277/1563 (17.7%) | 4.83 (3.51-6.41) Sum=7434 | 3.7 (3.3-4.2) | 453/4091 (11.1%) | 5.10 (3.83-6.55) Sum=20789 | 2.2 (2.0-2.4) | 1.71 (1.47 - 1.98) p=<.0001 | 1.65 (1.41 - 1.93) p=<.0001 | 0.0010 | 0.0011 |
|  | **2011-2015 vs. 2016-2020** | **2011-2015** | 277/1563 (17.7%) | 4.83 (3.51-6.41) Sum=7434 | 3.7 (3.3-4.2) | 453/4091 (11.1%) | 5.10 (3.83-6.55) Sum=20789 | 2.2 (2.0-2.4) | 1.71 (1.47 - 1.98) p=<.0001 | 1.65 (1.41 - 1.93) p=<.0001 |  |  |
|  |  | **2016-2020** | 284/1561 (18.2%) | 4.92 (3.50-6.47) Sum=7469 | 3.8 (3.4-4.3) | 447/4033 (11.1%) | 5.25 (3.92-6.61) Sum=20789 | 2.2 (2.0-2.4) | 1.76 (1.52 - 2.04) p=<.0001 | 1.63 (1.39 - 1.91) p=<.0001 | 0.76 | 0.68 |
| **30-64 years of age at inclusion** | **2006-2010 vs. 2016-2020** | **2006-2010** | 725/3682 (19.7%) | 4.23 (2.03-5.91) Sum=14772 | 4.9 (4.6-5.3) | 808/9410 (8.6%) | 5.30 (3.98-6.59) Sum=48825 | 1.7 (1.5-1.8) | 2.87 (2.59 - 3.17) p=<.0001 | 2.21 (1.97 - 2.49) p=<.0001 |  |  |
|  |  | **2016-2020** | 613/2879 (21.3%) | 4.34 (2.54-6.09) Sum=12023 | 5.1 (4.7-5.5) | 570/6559 (8.7%) | 5.37 (3.98-6.70) Sum=34478 | 1.7 (1.5-1.8) | 3.01 (2.68 - 3.37) p=<.0001 | 2.31 (2.03 - 2.64) p=<.0001 | 0.58 | 0.43 |
|  | **2006-2010 vs. 2011-2015** | **2006-2010** | 725/3682 (19.7%) | 4.23 (2.03-5.91) Sum=14772 | 4.9 (4.6-5.3) | 808/9410 (8.6%) | 5.30 (3.98-6.59) Sum=48825 | 1.7 (1.5-1.8) | 2.87 (2.59 - 3.17) p=<.0001 | 2.21 (1.97 - 2.49) p=<.0001 |  |  |
|  |  | **2011-2015** | 690/3357 (20.6%) | 4.29 (2.21-6.13) Sum=13852 | 5.0 (4.6-5.4) | 650/8086 (8.0%) | 5.37 (4.01-6.70) Sum=42603 | 1.5 (1.4-1.6) | 3.16 (2.84 - 3.52) p=<.0001 | 2.41 (2.13 - 2.73) p=<.0001 | 0.19 | 0.21 |
|  | **2011-2015 vs. 2016-2020** | **2011-2015** | 690/3357 (20.6%) | 4.29 (2.21-6.13) Sum=13852 | 5.0 (4.6-5.4) | 650/8086 (8.0%) | 5.37 (4.01-6.70) Sum=42603 | 1.5 (1.4-1.6) | 3.16 (2.84 - 3.52) p=<.0001 | 2.41 (2.13 - 2.73) p=<.0001 |  |  |
|  |  | **2016-2020** | 613/2879 (21.3%) | 4.34 (2.54-6.09) Sum=12023 | 5.1 (4.7-5.5) | 570/6559 (8.7%) | 5.37 (3.98-6.70) Sum=34478 | 1.7 (1.5-1.8) | 3.01 (2.68 - 3.37) p=<.0001 | 2.31 (2.03 - 2.64) p=<.0001 | 0.49 | 0.72 |
| **≥65 years of age at inclusion** | **2006-2010 vs. 2016-2020** | **2006-2010** | 1002/4456 (22.5%) | 2.95 (0.66-4.73) Sum=13073 | 7.7 (7.2-8.2) | 1526/10758 (14.2%) | 4.44 (3.19-5.85) Sum=46909 | 3.3 (3.1-3.4) | 2.27 (2.10 - 2.46) p=<.0001 | 1.68 (1.52 - 1.85) p=<.0001 |  |  |
|  |  | **2016-2020** | 1123/4725 (23.8%) | 3.23 (0.88-4.99) Sum=15135 | 7.4 (7.0-7.9) | 1543/10447 (14.8%) | 4.55 (3.26-6.07) Sum=47104 | 3.3 (3.1-3.4) | 2.21 (2.05 - 2.39) p=<.0001 | 1.70 (1.55 - 1.87) p=<.0001 | 0.55 | 0.32 |
|  | **2006-2010 vs. 2011-2015** | **2006-2010** | 1002/4456 (22.5%) | 2.95 (0.66-4.73) Sum=13073 | 7.7 (7.2-8.2) | 1526/10758 (14.2%) | 4.44 (3.19-5.85) Sum=46909 | 3.3 (3.1-3.4) | 2.27 (2.10 - 2.46) p=<.0001 | 1.68 (1.52 - 1.85) p=<.0001 |  |  |
|  |  | **2011-2015** | 1106/4923 (22.5%) | 3.12 (0.81-4.84) Sum=15108 | 7.3 (6.9-7.8) | 1598/11245 (14.2%) | 4.55 (3.24-6.05) Sum=50279 | 3.2 (3.0-3.3) | 2.23 (2.07 - 2.41) p=<.0001 | 1.70 (1.54 - 1.86) p=<.0001 | 0.71 | 0.39 |
|  | **2011-2015 vs. 2016-2020** | **2011-2015** | 1106/4923 (22.5%) | 3.12 (0.81-4.84) Sum=15108 | 7.3 (6.9-7.8) | 1598/11245 (14.2%) | 4.55 (3.24-6.05) Sum=50279 | 3.2 (3.0-3.3) | 2.23 (2.07 - 2.41) p=<.0001 | 1.70 (1.54 - 1.86) p=<.0001 |  |  |
|  |  | **2016-2020** | 1123/4725 (23.8%) | 3.23 (0.88-4.99) Sum=15135 | 7.4 (7.0-7.9) | 1543/10447 (14.8%) | 4.55 (3.26-6.07) Sum=47104 | 3.3 (3.1-3.4) | 2.21 (2.05 - 2.39) p=<.0001 | 1.70 (1.55 - 1.87) p=<.0001 | 0.81 | 0.91 |
| ***Male - population*** |  |  |  |  |  |  |  |  |  |  |  |  |
| **<30 years of age at inclusion** | **2006-2010 vs. 2016-2020** | **2006-2010** | 119/727 (16.4%) | 4.65 (3.47-5.98) Sum=3316 | 3.6 (3.0-4.3) | 119/2036 (5.8%) | 4.98 (3.87-6.29) Sum=10332 | 1.2 (1.0-1.4) | 3.09 (2.40 - 3.99) p=<.0001 | 2.96 (2.25 - 3.88) p=<.0001 |  |  |
|  |  | **2016-2020** | 139/894 (15.5%) | 4.96 (3.62-6.49) Sum=4317 | 3.2 (2.7-3.8) | 210/2386 (8.8%) | 5.29 (3.96-6.63) Sum=12497 | 1.7 (1.5-1.9) | 1.91 (1.54 - 2.37) p=<.0001 | 1.76 (1.40 - 2.21) p=<.0001 | 0.0043 | 0.0055 |
|  | **2006-2010 vs. 2011-2015** | **2006-2010** | 119/727 (16.4%) | 4.65 (3.47-5.98) Sum=3316 | 3.6 (3.0-4.3) | 119/2036 (5.8%) | 4.98 (3.87-6.29) Sum=10332 | 1.2 (1.0-1.4) | 3.09 (2.40 - 3.99) p=<.0001 | 2.96 (2.25 - 3.88) p=<.0001 |  |  |
|  |  | **2011-2015** | 134/824 (16.3%) | 4.80 (3.51-6.39) Sum=3907 | 3.4 (2.9-4.1) | 173/2229 (7.8%) | 5.15 (3.93-6.60) Sum=11495 | 1.5 (1.3-1.7) | 2.26 (1.81 - 2.83) p=<.0001 | 2.30 (1.81 - 2.92) p=<.0001 | 0.07 | 0.07 |
|  | **2011-2015 vs. 2016-2020** | **2011-2015** | 134/824 (16.3%) | 4.80 (3.51-6.39) Sum=3907 | 3.4 (2.9-4.1) | 173/2229 (7.8%) | 5.15 (3.93-6.60) Sum=11495 | 1.5 (1.3-1.7) | 2.26 (1.81 - 2.83) p=<.0001 | 2.30 (1.81 - 2.92) p=<.0001 |  |  |
|  |  | **2016-2020** | 139/894 (15.5%) | 4.96 (3.62-6.49) Sum=4317 | 3.2 (2.7-3.8) | 210/2386 (8.8%) | 5.29 (3.96-6.63) Sum=12497 | 1.7 (1.5-1.9) | 1.91 (1.54 - 2.37) p=<.0001 | 1.76 (1.40 - 2.21) p=<.0001 | 0.28 | 0.31 |
| **30-64 years of age at inclusion** | **2006-2010 vs. 2016-2020** | **2006-2010** | 393/2112 (18.6%) | 4.25 (1.81-5.94) Sum=8438 | 4.7 (4.2-5.1) | 378/5606 (6.7%) | 5.39 (4.10-6.63) Sum=29489 | 1.3 (1.2-1.4) | 3.48 (3.02 - 4.01) p=<.0001 | 2.56 (2.16 - 3.03) p=<.0001 |  |  |
|  |  | **2016-2020** | 351/1721 (20.4%) | 4.32 (2.40-6.07) Sum=7158 | 4.9 (4.4-5.4) | 286/4138 (6.9%) | 5.38 (4.00-6.64) Sum=21782 | 1.3 (1.2-1.5) | 3.63 (3.11 - 4.25) p=<.0001 | 2.75 (2.29 - 3.31) p=<.0001 | 0.76 | 0.80 |
|  | **2006-2010 vs. 2011-2015** | **2006-2010** | 393/2112 (18.6%) | 4.25 (1.81-5.94) Sum=8438 | 4.7 (4.2-5.1) | 378/5606 (6.7%) | 5.39 (4.10-6.63) Sum=29489 | 1.3 (1.2-1.4) | 3.48 (3.02 - 4.01) p=<.0001 | 2.56 (2.16 - 3.03) p=<.0001 |  |  |
|  |  | **2011-2015** | 375/1962 (19.1%) | 4.38 (2.17-6.15) Sum=8136 | 4.6 (4.2-5.1) | 313/4928 (6.4%) | 5.39 (4.05-6.71) Sum=26160 | 1.2 (1.1-1.3) | 3.71 (3.20 - 4.32) p=<.0001 | 2.85 (2.39 - 3.41) p=<.0001 | 0.54 | 0.69 |
|  | **2011-2015 vs. 2016-2020** | **2011-2015** | 375/1962 (19.1%) | 4.38 (2.17-6.15) Sum=8136 | 4.6 (4.2-5.1) | 313/4928 (6.4%) | 5.39 (4.05-6.71) Sum=26160 | 1.2 (1.1-1.3) | 3.71 (3.20 - 4.32) p=<.0001 | 2.85 (2.39 - 3.41) p=<.0001 |  |  |
|  |  | **2016-2020** | 351/1721 (20.4%) | 4.32 (2.40-6.07) Sum=7158 | 4.9 (4.4-5.4) | 286/4138 (6.9%) | 5.38 (4.00-6.64) Sum=21782 | 1.3 (1.2-1.5) | 3.63 (3.11 - 4.25) p=<.0001 | 2.75 (2.29 - 3.31) p=<.0001 | 0.79 | 0.93 |
| **≥65 years of age at inclusion** | **2006-2010 vs. 2016-2020** | **2006-2010** | 507/2472 (20.5%) | 3.16 (0.76-4.85) Sum=7523 | 6.7 (6.2-7.4) | 751/6222 (12.1%) | 4.50 (3.24-5.88) Sum=27506 | 2.7 (2.5-2.9) | 2.39 (2.14 - 2.68) p=<.0001 | 1.72 (1.50 - 1.97) p=<.0001 |  |  |
|  |  | **2016-2020** | 639/2930 (21.8%) | 3.33 (1.02-5.08) Sum=9646 | 6.6 (6.1-7.2) | 899/6805 (13.2%) | 4.55 (3.29-6.07) Sum=30767 | 2.9 (2.7-3.1) | 2.22 (2.00 - 2.46) p=<.0001 | 1.67 (1.48 - 1.88) p=<.0001 | 0.30 | 0.21 |
|  | **2006-2010 vs. 2011-2015** | **2006-2010** | 507/2472 (20.5%) | 3.16 (0.76-4.85) Sum=7523 | 6.7 (6.2-7.4) | 751/6222 (12.1%) | 4.50 (3.24-5.88) Sum=27506 | 2.7 (2.5-2.9) | 2.39 (2.14 - 2.68) p=<.0001 | 1.72 (1.50 - 1.97) p=<.0001 |  |  |
|  |  | **2011-2015** | 598/2906 (20.6%) | 3.24 (0.87-4.96) Sum=9190 | 6.5 (6.0-7.1) | 868/6964 (12.5%) | 4.55 (3.25-6.05) Sum=31282 | 2.8 (2.6-3.0) | 2.27 (2.05 - 2.52) p=<.0001 | 1.63 (1.44 - 1.85) p=<.0001 | 0.53 | 0.28 |
|  | **2011-2015 vs. 2016-2020** | **2011-2015** | 598/2906 (20.6%) | 3.24 (0.87-4.96) Sum=9190 | 6.5 (6.0-7.1) | 868/6964 (12.5%) | 4.55 (3.25-6.05) Sum=31282 | 2.8 (2.6-3.0) | 2.27 (2.05 - 2.52) p=<.0001 | 1.63 (1.44 - 1.85) p=<.0001 |  |  |
|  |  | **2016-2020** | 639/2930 (21.8%) | 3.33 (1.02-5.08) Sum=9646 | 6.6 (6.1-7.2) | 899/6805 (13.2%) | 4.55 (3.29-6.07) Sum=30767 | 2.9 (2.7-3.1) | 2.22 (2.00 - 2.46) p=<.0001 | 1.67 (1.48 - 1.88) p=<.0001 | 0.68 | 0.90 |
| ***Female - population*** |  |  |  |  |  |  |  |  |  |  |  |  |
| **<30 years of age at inclusion** | **2006-2010 vs. 2016-2020** | **2006-2010** | 140/696 (20.1%) | 4.68 (3.54-6.11) Sum=3236 | 4.3 (3.6-5.1) | 195/1844 (10.6%) | 5.07 (3.93-6.38) Sum=9399 | 2.1 (1.8-2.4) | 2.10 (1.69 - 2.60) p=<.0001 | 1.98 (1.57 - 2.49) p=<.0001 |  |  |
|  |  | **2016-2020** | 145/667 (21.7%) | 4.82 (3.41-6.41) Sum=3151 | 4.6 (3.9-5.4) | 237/1647 (14.4%) | 5.17 (3.79-6.54) Sum=8292 | 2.9 (2.5-3.2) | 1.60 (1.30 - 1.97) p=<.0001 | 1.53 (1.23 - 1.91) p=0.0001 | 0.09 | 0.17 |
|  | **2006-2010 vs. 2011-2015** | **2006-2010** | 140/696 (20.1%) | 4.68 (3.54-6.11) Sum=3236 | 4.3 (3.6-5.1) | 195/1844 (10.6%) | 5.07 (3.93-6.38) Sum=9399 | 2.1 (1.8-2.4) | 2.10 (1.69 - 2.60) p=<.0001 | 1.98 (1.57 - 2.49) p=<.0001 |  |  |
|  |  | **2011-2015** | 143/739 (19.4%) | 4.85 (3.53-6.41) Sum=3527 | 4.1 (3.4-4.8) | 280/1862 (15.0%) | 5.04 (3.72-6.48) Sum=9294 | 3.0 (2.7-3.4) | 1.34 (1.10 - 1.65) p=0.0039 | 1.29 (1.04 - 1.60) p=0.019 | 0.0036 | 0.0047 |
|  | **2011-2015 vs. 2016-2020** | **2011-2015** | 143/739 (19.4%) | 4.85 (3.53-6.41) Sum=3527 | 4.1 (3.4-4.8) | 280/1862 (15.0%) | 5.04 (3.72-6.48) Sum=9294 | 3.0 (2.7-3.4) | 1.34 (1.10 - 1.65) p=0.0039 | 1.29 (1.04 - 1.60) p=0.019 |  |  |
|  |  | **2016-2020** | 145/667 (21.7%) | 4.82 (3.41-6.41) Sum=3151 | 4.6 (3.9-5.4) | 237/1647 (14.4%) | 5.17 (3.79-6.54) Sum=8292 | 2.9 (2.5-3.2) | 1.60 (1.30 - 1.97) p=<.0001 | 1.53 (1.23 - 1.91) p=0.0001 | 0.22 | 0.19 |
| **30-64 years of age at inclusion** | **2006-2010 vs. 2016-2020** | **2006-2010** | 332/1570 (21.1%) | 4.21 (2.33-5.84) Sum=6335 | 5.2 (4.7-5.8) | 430/3804 (11.3%) | 5.13 (3.81-6.54) Sum=19336 | 2.2 (2.0-2.4) | 2.30 (1.99 - 2.66) p=<.0001 | 1.93 (1.64 - 2.28) p=<.0001 |  |  |
|  |  | **2016-2020** | 262/1158 (22.6%) | 4.39 (2.59-6.13) Sum=4865 | 5.4 (4.8-6.1) | 284/2421 (11.7%) | 5.36 (3.94-6.76) Sum=12695 | 2.2 (2.0-2.5) | 2.36 (1.99 - 2.79) p=<.0001 | 1.94 (1.61 - 2.35) p=<.0001 | 0.84 | 0.58 |
|  | **2006-2010 vs. 2011-2015** | **2006-2010** | 332/1570 (21.1%) | 4.21 (2.33-5.84) Sum=6335 | 5.2 (4.7-5.8) | 430/3804 (11.3%) | 5.13 (3.81-6.54) Sum=19336 | 2.2 (2.0-2.4) | 2.30 (1.99 - 2.66) p=<.0001 | 1.93 (1.64 - 2.28) p=<.0001 |  |  |
|  |  | **2011-2015** | 315/1395 (22.6%) | 4.20 (2.23-6.12) Sum=5716 | 5.5 (4.9-6.2) | 337/3158 (10.7%) | 5.34 (3.93-6.65) Sum=16442 | 2.0 (1.8-2.3) | 2.62 (2.24 - 3.05) p=<.0001 | 2.04 (1.71 - 2.44) p=<.0001 | 0.22 | 0.22 |
|  | **2011-2015 vs. 2016-2020** | **2011-2015** | 315/1395 (22.6%) | 4.20 (2.23-6.12) Sum=5716 | 5.5 (4.9-6.2) | 337/3158 (10.7%) | 5.34 (3.93-6.65) Sum=16442 | 2.0 (1.8-2.3) | 2.62 (2.24 - 3.05) p=<.0001 | 2.04 (1.71 - 2.44) p=<.0001 |  |  |
|  |  | **2016-2020** | 262/1158 (22.6%) | 4.39 (2.59-6.13) Sum=4865 | 5.4 (4.8-6.1) | 284/2421 (11.7%) | 5.36 (3.94-6.76) Sum=12695 | 2.2 (2.0-2.5) | 2.36 (1.99 - 2.79) p=<.0001 | 1.94 (1.61 - 2.35) p=<.0001 | 0.35 | 0.59 |
| **≥65 years of age at inclusion** | **2006-2010 vs. 2016-2020** | **2006-2010** | 495/1984 (24.9%) | 2.57 (0.56-4.58) Sum=5550 | 8.9 (8.2-9.7) | 775/4536 (17.1%) | 4.32 (3.12-5.78) Sum=19403 | 4.0 (3.7-4.3) | 2.14 (1.91 - 2.40) p=<.0001 | 1.64 (1.43 - 1.89) p=<.0001 |  |  |
|  |  | **2016-2020** | 484/1795 (27.0%) | 3.07 (0.73-4.84) Sum=5489 | 8.8 (8.0-9.6) | 644/3642 (17.7%) | 4.54 (3.20-6.05) Sum=16337 | 3.9 (3.6-4.3) | 2.18 (1.93 - 2.45) p=<.0001 | 1.78 (1.54 - 2.07) p=<.0001 | 0.93 | 0.84 |
|  | **2006-2010 vs. 2011-2015** | **2006-2010** | 495/1984 (24.9%) | 2.57 (0.56-4.58) Sum=5550 | 8.9 (8.2-9.7) | 775/4536 (17.1%) | 4.32 (3.12-5.78) Sum=19403 | 4.0 (3.7-4.3) | 2.14 (1.91 - 2.40) p=<.0001 | 1.64 (1.43 - 1.89) p=<.0001 |  |  |
|  |  | **2011-2015** | 508/2017 (25.2%) | 2.93 (0.71-4.65) Sum=5918 | 8.6 (7.9-9.4) | 730/4281 (17.1%) | 4.54 (3.20-6.05) Sum=18997 | 3.8 (3.6-4.1) | 2.16 (1.93 - 2.42) p=<.0001 | 1.79 (1.56 - 2.05) p=<.0001 | 0.98 | 0.81 |
|  | **2011-2015 vs. 2016-2020** | **2011-2015** | 508/2017 (25.2%) | 2.93 (0.71-4.65) Sum=5918 | 8.6 (7.9-9.4) | 730/4281 (17.1%) | 4.54 (3.20-6.05) Sum=18997 | 3.8 (3.6-4.1) | 2.16 (1.93 - 2.42) p=<.0001 | 1.79 (1.56 - 2.05) p=<.0001 |  |  |
|  |  | **2016-2020** | 484/1795 (27.0%) | 3.07 (0.73-4.84) Sum=5489 | 8.8 (8.0-9.6) | 644/3642 (17.7%) | 4.54 (3.20-6.05) Sum=16337 | 3.9 (3.6-4.3) | 2.18 (1.93 - 2.45) p=<.0001 | 1.78 (1.54 - 2.07) p=<.0001 | 0.95 | 0.97 |
| Confidence interval for unadjusted event rates per 100 person years are obtained from exact Poisson confidence limits. Cox regression was used for time to any event presented by HR. * Adjusted for Age at inclusion, Sex, Stroke, TBI, Dementia, Brain tumour, Brain infection, Diabetes, Cardiovascular diseases, Intellectual disability, Cancer. The p-value for interaction was calculated between study group and each pairwise time period. | | | | | | | | | | | | |

### **eTable 4A. Unadjusted, and fully adjusted HR, CIs, p-values for first specialist psychiatric care from Cox regression in cases vs controls during periods 2006-2010, 2011-2015 and 2016-2020 overall and by sex (FAS population)**

|  | | | **Case** | | | **Control** | | | **Case vs. Control** | | **Interaction** | |
| --- | --- | --- | --- | --- | --- | --- | --- | --- | --- | --- | --- | --- |
| **Subgroup** | **Period** | **Period** | **n/N (%) events** | **Follow-up time Median (IQR) Sum** | **Event rate (95% CI) per 100 person- years** | **n/N (%) events** | **Follow-up time Median (IQR) Sum** | **Event rate (95% CI) per 100 person- years** | **Hazard ratio (95% CI) p-value** | *** Hazard ratio (95% CI) p-value** | **p-value** | *** p-value** |
| ***Overall - population*** |  |  |  |  |  |  |  |  |  |  |  |  |
|  | **2006-2010 vs. 2016-2020** | **2006-2010** | 230/9561 (2.4%) | 4.25 (2.52-5.88) Sum=39065 | 0.59 (0.52-0.67) | 210/24048 (0.9%) | 5.10 (3.84-6.45) Sum=121658 | 0.17 (0.15-0.20) | 3.35 (2.78 - 4.05) p=<.0001 | 2.97 (2.41 - 3.66) p=<.0001 |  |  |
|  |  | **2016-2020** | 148/9165 (1.6%) | 4.45 (3.05-6.12) Sum=39485 | 0.37 (0.32-0.44) | 103/21039 (0.5%) | 5.20 (3.90-6.58) Sum=108447 | 0.09 (0.08-0.12) | 3.87 (3.01 - 4.98) p=<.0001 | 3.54 (2.69 - 4.65) p=<.0001 | 0.35 | 0.20 |
|  | **2006-2010 vs. 2011-2015** | **2006-2010** | 230/9561 (2.4%) | 4.25 (2.52-5.88) Sum=39065 | 0.59 (0.52-0.67) | 210/24048 (0.9%) | 5.10 (3.84-6.45) Sum=121658 | 0.17 (0.15-0.20) | 3.35 (2.78 - 4.05) p=<.0001 | 2.97 (2.41 - 3.66) p=<.0001 |  |  |
|  |  | **2011-2015** | 203/9843 (2.1%) | 4.35 (2.91-6.04) Sum=41545 | 0.49 (0.42-0.56) | 197/23422 (0.8%) | 5.17 (3.87-6.55) Sum=119889 | 0.16 (0.14-0.19) | 2.95 (2.42 - 3.58) p=<.0001 | 2.68 (2.16 - 3.32) p=<.0001 | 0.33 | 0.32 |
|  | **2011-2015 vs. 2016-2020** | **2011-2015** | 203/9843 (2.1%) | 4.35 (2.91-6.04) Sum=41545 | 0.49 (0.42-0.56) | 197/23422 (0.8%) | 5.17 (3.87-6.55) Sum=119889 | 0.16 (0.14-0.19) | 2.95 (2.42 - 3.58) p=<.0001 | 2.68 (2.16 - 3.32) p=<.0001 |  |  |
|  |  | **2016-2020** | 148/9165 (1.6%) | 4.45 (3.05-6.12) Sum=39485 | 0.37 (0.32-0.44) | 103/21039 (0.5%) | 5.20 (3.90-6.58) Sum=108447 | 0.09 (0.08-0.12) | 3.87 (3.01 - 4.98) p=<.0001 | 3.54 (2.69 - 4.65) p=<.0001 | 0.08 | 0.049 |
| ***Male - population*** |  |  |  |  |  |  |  |  |  |  |  |  |
|  | **2006-2010 vs. 2016-2020** | **2006-2010** | 105/5311 (2.0%) | 4.25 (2.52-5.91) Sum=21738 | 0.48 (0.40-0.58) | 108/13864 (0.8%) | 5.13 (3.85-6.42) Sum=70137 | 0.15 (0.13-0.19) | 3.09 (2.36 - 4.04) p=<.0001 | 2.63 (1.94 - 3.57) p=<.0001 |  |  |
|  |  | **2016-2020** | 86/5545 (1.6%) | 4.39 (3.03-6.07) Sum=23674 | 0.36 (0.29-0.45) | 68/13329 (0.5%) | 5.14 (3.87-6.53) Sum=68074 | 0.10 (0.08-0.13) | 3.54 (2.58 - 4.87) p=<.0001 | 3.16 (2.23 - 4.49) p=<.0001 | 0.48 | 0.26 |
|  | **2006-2010 vs. 2011-2015** | **2006-2010** | 105/5311 (2.0%) | 4.25 (2.52-5.91) Sum=21738 | 0.48 (0.40-0.58) | 108/13864 (0.8%) | 5.13 (3.85-6.42) Sum=70137 | 0.15 (0.13-0.19) | 3.09 (2.36 - 4.04) p=<.0001 | 2.63 (1.94 - 3.57) p=<.0001 |  |  |
|  |  | **2011-2015** | 104/5692 (1.8%) | 4.34 (2.82-6.01) Sum=23886 | 0.44 (0.36-0.53) | 110/14121 (0.8%) | 5.13 (3.84-6.54) Sum=71887 | 0.15 (0.13-0.18) | 2.80 (2.14 - 3.67) p=<.0001 | 2.53 (1.88 - 3.43) p=<.0001 | 0.62 | 0.70 |
|  | **2011-2015 vs. 2016-2020** | **2011-2015** | 104/5692 (1.8%) | 4.34 (2.82-6.01) Sum=23886 | 0.44 (0.36-0.53) | 110/14121 (0.8%) | 5.13 (3.84-6.54) Sum=71887 | 0.15 (0.13-0.18) | 2.80 (2.14 - 3.67) p=<.0001 | 2.53 (1.88 - 3.43) p=<.0001 |  |  |
|  |  | **2016-2020** | 86/5545 (1.6%) | 4.39 (3.03-6.07) Sum=23674 | 0.36 (0.29-0.45) | 68/13329 (0.5%) | 5.14 (3.87-6.53) Sum=68074 | 0.10 (0.08-0.13) | 3.54 (2.58 - 4.87) p=<.0001 | 3.16 (2.23 - 4.49) p=<.0001 | 0.25 | 0.17 |
| ***Female - population*** |  |  |  |  |  |  |  |  |  |  |  |  |
|  | **2006-2010 vs. 2016-2020** | **2006-2010** | 125/4250 (2.9%) | 4.23 (2.51-5.86) Sum=17327 | 0.72 (0.60-0.86) | 102/10184 (1.0%) | 5.07 (3.83-6.49) Sum=51521 | 0.20 (0.16-0.24) | 3.58 (2.75 - 4.65) p=<.0001 | 3.31 (2.48 - 4.41) p=<.0001 |  |  |
|  |  | **2016-2020** | 62/3620 (1.7%) | 4.56 (3.10-6.21) Sum=15812 | 0.39 (0.30-0.50) | 35/7710 (0.5%) | 5.31 (3.97-6.69) Sum=40373 | 0.09 (0.06-0.12) | 4.47 (2.95 - 6.77) p=<.0001 | 4.25 (2.74 - 6.60) p=<.0001 | 0.38 | 0.31 |
|  | **2006-2010 vs. 2011-2015** | **2006-2010** | 125/4250 (2.9%) | 4.23 (2.51-5.86) Sum=17327 | 0.72 (0.60-0.86) | 102/10184 (1.0%) | 5.07 (3.83-6.49) Sum=51521 | 0.20 (0.16-0.24) | 3.58 (2.75 - 4.65) p=<.0001 | 3.31 (2.48 - 4.41) p=<.0001 |  |  |
|  |  | **2011-2015** | 99/4151 (2.4%) | 4.36 (3.01-6.07) Sum=17658 | 0.56 (0.46-0.68) | 87/9301 (0.9%) | 5.23 (3.90-6.57) Sum=48002 | 0.18 (0.15-0.22) | 3.08 (2.31 - 4.11) p=<.0001 | 2.82 (2.06 - 3.85) p=<.0001 | 0.41 | 0.37 |
|  | **2011-2015 vs. 2016-2020** | **2011-2015** | 99/4151 (2.4%) | 4.36 (3.01-6.07) Sum=17658 | 0.56 (0.46-0.68) | 87/9301 (0.9%) | 5.23 (3.90-6.57) Sum=48002 | 0.18 (0.15-0.22) | 3.08 (2.31 - 4.11) p=<.0001 | 2.82 (2.06 - 3.85) p=<.0001 |  |  |
|  |  | **2016-2020** | 62/3620 (1.7%) | 4.56 (3.10-6.21) Sum=15812 | 0.39 (0.30-0.50) | 35/7710 (0.5%) | 5.31 (3.97-6.69) Sum=40373 | 0.09 (0.06-0.12) | 4.47 (2.95 - 6.77) p=<.0001 | 4.25 (2.74 - 6.60) p=<.0001 | 0.14 | 0.11 |
| Confidence interval for unadjusted event rates per 100 person years are obtained from exact Poisson confidence limits. Cox regression was used for time to any event presented by HR. * Adjusted for Age at inclusion, Sex, Stroke, TBI, Dementia, Brain tumour, Brain infection, Diabetes, Cardiovascular diseases, Intellectual disability, Cancer, Specialist psychiatric care (before date of inclusion). The p-value for interaction was calculated between study group and each pairwise time period. | | | | | | | | | | | | |

### **eTable 4B. Unadjusted, and fully adjusted HR, CIs, p-values for first specialist psychiatric care from Cox regression in cases vs controls during periods 2006-2010, 2011-2015 and 2016-2020 for each subgroup overall and by sex (FAS population)**

|  | | | **Case** | | | **Control** | | | **Case vs. Control** | | **Interaction** | |
| --- | --- | --- | --- | --- | --- | --- | --- | --- | --- | --- | --- | --- |
| **Subgroup** | **Period** | **Period** | **n/N (%) events** | **Follow-up time Median (IQR) Sum** | **Event rate (95% CI) per 100 person- years** | **n/N (%) events** | **Follow-up time Median (IQR) Sum** | **Event rate (95% CI) per 100 person- years** | **Hazard ratio (95% CI) p-value** | *** Hazard ratio (95% CI) p-value** | **p-value** | *** p-value** |
| ***Overall - population*** |  |  |  |  |  |  |  |  |  |  |  |  |
| **<30 years of age at inclusion** | **2006-2010 vs. 2016-2020** | **2006-2010** | 78/1423 (5.5%) | 4.90 (3.81-6.30) Sum=7089 | 1.10 (0.87-1.37) | 57/3880 (1.5%) | 5.19 (4.08-6.49) Sum=20402 | 0.28 (0.21-0.36) | 3.91 (2.78 - 5.51) p=<.0001 |  |  |  |
|  |  | **2016-2020** | 73/1561 (4.7%) | 5.28 (3.97-6.66) Sum=8126 | 0.90 (0.70-1.13) | 52/4033 (1.3%) | 5.55 (4.25-6.74) Sum=22005 | 0.24 (0.18-0.31) | 3.77 (2.64 - 5.39) p=<.0001 |  | 0.89 |  |
|  | **2006-2010 vs. 2011-2015** | **2006-2010** | 78/1423 (5.5%) | 4.90 (3.81-6.30) Sum=7089 | 1.10 (0.87-1.37) | 57/3880 (1.5%) | 5.19 (4.08-6.49) Sum=20402 | 0.28 (0.21-0.36) | 3.91 (2.78 - 5.51) p=<.0001 |  |  |  |
|  |  | **2011-2015** | 67/1563 (4.3%) | 5.21 (3.93-6.60) Sum=8116 | 0.83 (0.64-1.05) | 77/4091 (1.9%) | 5.31 (4.08-6.72) Sum=21868 | 0.35 (0.28-0.44) | 2.34 (1.69 - 3.25) p=<.0001 |  | 0.033 |  |
|  | **2011-2015 vs. 2016-2020** | **2011-2015** | 67/1563 (4.3%) | 5.21 (3.93-6.60) Sum=8116 | 0.83 (0.64-1.05) | 77/4091 (1.9%) | 5.31 (4.08-6.72) Sum=21868 | 0.35 (0.28-0.44) | 2.34 (1.69 - 3.25) p=<.0001 |  |  |  |
|  |  | **2016-2020** | 73/1561 (4.7%) | 5.28 (3.97-6.66) Sum=8126 | 0.90 (0.70-1.13) | 52/4033 (1.3%) | 5.55 (4.25-6.74) Sum=22005 | 0.24 (0.18-0.31) | 3.77 (2.64 - 5.39) p=<.0001 |  | 0.051 |  |
| **30-64 years of age at inclusion** | **2006-2010 vs. 2016-2020** | **2006-2010** | 109/3682 (3.0%) | 4.75 (3.30-6.26) Sum=16678 | 0.65 (0.54-0.79) | 86/9410 (0.9%) | 5.52 (4.22-6.70) Sum=51057 | 0.17 (0.13-0.21) | 3.80 (2.86 - 5.05) p=<.0001 |  |  |  |
|  |  | **2016-2020** | 48/2879 (1.7%) | 4.95 (3.48-6.46) Sum=13707 | 0.35 (0.26-0.46) | 27/6559 (0.4%) | 5.60 (4.23-6.85) Sum=36060 | 0.07 (0.05-0.11) | 4.54 (2.83 - 7.27) p=<.0001 |  | 0.49 |  |
|  | **2006-2010 vs. 2011-2015** | **2006-2010** | 109/3682 (3.0%) | 4.75 (3.30-6.26) Sum=16678 | 0.65 (0.54-0.79) | 86/9410 (0.9%) | 5.52 (4.22-6.70) Sum=51057 | 0.17 (0.13-0.21) | 3.80 (2.86 - 5.05) p=<.0001 |  |  |  |
|  |  | **2011-2015** | 86/3357 (2.6%) | 4.89 (3.40-6.45) Sum=15793 | 0.54 (0.44-0.67) | 62/8086 (0.8%) | 5.58 (4.23-6.81) Sum=44323 | 0.14 (0.11-0.18) | 3.85 (2.77 - 5.33) p=<.0001 |  | 0.98 |  |
|  | **2011-2015 vs. 2016-2020** | **2011-2015** | 86/3357 (2.6%) | 4.89 (3.40-6.45) Sum=15793 | 0.54 (0.44-0.67) | 62/8086 (0.8%) | 5.58 (4.23-6.81) Sum=44323 | 0.14 (0.11-0.18) | 3.85 (2.77 - 5.33) p=<.0001 |  |  |  |
|  |  | **2016-2020** | 48/2879 (1.7%) | 4.95 (3.48-6.46) Sum=13707 | 0.35 (0.26-0.46) | 27/6559 (0.4%) | 5.60 (4.23-6.85) Sum=36060 | 0.07 (0.05-0.11) | 4.54 (2.83 - 7.27) p=<.0001 |  | 0.53 |  |
| **≥65 years of age at inclusion** | **2006-2010 vs. 2016-2020** | **2006-2010** | 43/4456 (1.0%) | 3.53 (1.23-5.22) Sum=15297 | 0.28 (0.20-0.38) | 67/10758 (0.6%) | 4.71 (3.48-6.09) Sum=50199 | 0.13 (0.10-0.17) | 2.07 (1.41 - 3.05) p=0.0002 |  |  |  |
|  |  | **2016-2020** | 27/4725 (0.6%) | 3.84 (1.75-5.49) Sum=17652 | 0.15 (0.10-0.22) | 24/10447 (0.2%) | 4.85 (3.59-6.28) Sum=50382 | 0.05 (0.03-0.07) | 3.17 (1.83 - 5.50) p=<.0001 |  | 0.21 |  |
|  | **2006-2010 vs. 2011-2015** | **2006-2010** | 43/4456 (1.0%) | 3.53 (1.23-5.22) Sum=15297 | 0.28 (0.20-0.38) | 67/10758 (0.6%) | 4.71 (3.48-6.09) Sum=50199 | 0.13 (0.10-0.17) | 2.07 (1.41 - 3.05) p=0.0002 |  |  |  |
|  |  | **2011-2015** | 50/4923 (1.0%) | 3.70 (1.45-5.34) Sum=17635 | 0.28 (0.21-0.37) | 58/11245 (0.5%) | 4.83 (3.56-6.26) Sum=53698 | 0.11 (0.08-0.14) | 2.57 (1.76 - 3.76) p=<.0001 |  | 0.42 |  |
|  | **2011-2015 vs. 2016-2020** | **2011-2015** | 50/4923 (1.0%) | 3.70 (1.45-5.34) Sum=17635 | 0.28 (0.21-0.37) | 58/11245 (0.5%) | 4.83 (3.56-6.26) Sum=53698 | 0.11 (0.08-0.14) | 2.57 (1.76 - 3.76) p=<.0001 |  |  |  |
|  |  | **2016-2020** | 27/4725 (0.6%) | 3.84 (1.75-5.49) Sum=17652 | 0.15 (0.10-0.22) | 24/10447 (0.2%) | 4.85 (3.59-6.28) Sum=50382 | 0.05 (0.03-0.07) | 3.17 (1.83 - 5.50) p=<.0001 |  | 0.55 |  |
| ***Male - population*** |  |  |  |  |  |  |  |  |  |  |  |  |
| **<30 years of age at inclusion** | **2006-2010 vs. 2016-2020** | **2006-2010** | 36/727 (5.0%) | 4.85 (3.76-6.25) Sum=3579 | 1.01 (0.70-1.39) | 19/2036 (0.9%) | 5.10 (4.02-6.40) Sum=10605 | 0.18 (0.11-0.28) | 5.60 (3.21 - 9.76) p=<.0001 |  |  |  |
|  |  | **2016-2020** | 37/894 (4.1%) | 5.29 (3.96-6.63) Sum=4637 | 0.80 (0.56-1.10) | 34/2386 (1.4%) | 5.57 (4.25-6.74) Sum=12994 | 0.26 (0.18-0.37) | 3.03 (1.90 - 4.83) p=<.0001 |  | 0.10 |  |
|  | **2006-2010 vs. 2011-2015** | **2006-2010** | 36/727 (5.0%) | 4.85 (3.76-6.25) Sum=3579 | 1.01 (0.70-1.39) | 19/2036 (0.9%) | 5.10 (4.02-6.40) Sum=10605 | 0.18 (0.11-0.28) | 5.60 (3.21 - 9.76) p=<.0001 |  |  |  |
|  |  | **2011-2015** | 29/824 (3.5%) | 5.13 (3.94-6.57) Sum=4252 | 0.68 (0.46-0.98) | 33/2229 (1.5%) | 5.28 (4.08-6.67) Sum=11899 | 0.28 (0.19-0.39) | 2.47 (1.50 - 4.07) p=0.0004 |  | 0.031 |  |
|  | **2011-2015 vs. 2016-2020** | **2011-2015** | 29/824 (3.5%) | 5.13 (3.94-6.57) Sum=4252 | 0.68 (0.46-0.98) | 33/2229 (1.5%) | 5.28 (4.08-6.67) Sum=11899 | 0.28 (0.19-0.39) | 2.47 (1.50 - 4.07) p=0.0004 |  |  |  |
|  |  | **2016-2020** | 37/894 (4.1%) | 5.29 (3.96-6.63) Sum=4637 | 0.80 (0.56-1.10) | 34/2386 (1.4%) | 5.57 (4.25-6.74) Sum=12994 | 0.26 (0.18-0.37) | 3.03 (1.90 - 4.83) p=<.0001 |  | 0.54 |  |
| **30-64 years of age at inclusion** | **2006-2010 vs. 2016-2020** | **2006-2010** | 47/2112 (2.2%) | 4.76 (3.23-6.28) Sum=9547 | 0.49 (0.36-0.65) | 45/5606 (0.8%) | 5.55 (4.25-6.70) Sum=30525 | 0.15 (0.11-0.20) | 3.27 (2.17 - 4.93) p=<.0001 |  |  |  |
|  |  | **2016-2020** | 33/1721 (1.9%) | 4.88 (3.35-6.35) Sum=8042 | 0.41 (0.28-0.58) | 20/4138 (0.5%) | 5.57 (4.19-6.78) Sum=22538 | 0.09 (0.05-0.14) | 4.40 (2.53 - 7.67) p=<.0001 |  | 0.35 |  |
|  | **2006-2010 vs. 2011-2015** | **2006-2010** | 47/2112 (2.2%) | 4.76 (3.23-6.28) Sum=9547 | 0.49 (0.36-0.65) | 45/5606 (0.8%) | 5.55 (4.25-6.70) Sum=30525 | 0.15 (0.11-0.20) | 3.27 (2.17 - 4.93) p=<.0001 |  |  |  |
|  |  | **2011-2015** | 44/1962 (2.2%) | 4.86 (3.33-6.44) Sum=9121 | 0.48 (0.35-0.65) | 43/4928 (0.9%) | 5.56 (4.25-6.80) Sum=26945 | 0.16 (0.12-0.21) | 2.97 (1.95 - 4.52) p=<.0001 |  | 0.75 |  |
|  | **2011-2015 vs. 2016-2020** | **2011-2015** | 44/1962 (2.2%) | 4.86 (3.33-6.44) Sum=9121 | 0.48 (0.35-0.65) | 43/4928 (0.9%) | 5.56 (4.25-6.80) Sum=26945 | 0.16 (0.12-0.21) | 2.97 (1.95 - 4.52) p=<.0001 |  |  |  |
|  |  | **2016-2020** | 33/1721 (1.9%) | 4.88 (3.35-6.35) Sum=8042 | 0.41 (0.28-0.58) | 20/4138 (0.5%) | 5.57 (4.19-6.78) Sum=22538 | 0.09 (0.05-0.14) | 4.40 (2.53 - 7.67) p=<.0001 |  | 0.23 |  |
| **≥65 years of age at inclusion** | **2006-2010 vs. 2016-2020** | **2006-2010** | 22/2472 (0.9%) | 3.59 (1.31-5.25) Sum=8613 | 0.26 (0.16-0.39) | 44/6222 (0.7%) | 4.72 (3.47-6.09) Sum=29007 | 0.15 (0.11-0.20) | 1.67 (1.00 - 2.79) p=0.050 |  |  |  |
|  |  | **2016-2020** | 16/2930 (0.5%) | 3.85 (1.86-5.47) Sum=10995 | 0.15 (0.08-0.24) | 14/6805 (0.2%) | 4.82 (3.56-6.22) Sum=32542 | 0.04 (0.02-0.07) | 3.37 (1.64 - 6.91) p=0.0009 |  | 0.12 |  |
|  | **2006-2010 vs. 2011-2015** | **2006-2010** | 22/2472 (0.9%) | 3.59 (1.31-5.25) Sum=8613 | 0.26 (0.16-0.39) | 44/6222 (0.7%) | 4.72 (3.47-6.09) Sum=29007 | 0.15 (0.11-0.20) | 1.67 (1.00 - 2.79) p=0.050 |  |  |  |
|  |  | **2011-2015** | 31/2906 (1.1%) | 3.74 (1.49-5.40) Sum=10513 | 0.29 (0.20-0.42) | 34/6964 (0.5%) | 4.81 (3.54-6.20) Sum=33043 | 0.10 (0.07-0.14) | 2.78 (1.71 - 4.53) p=<.0001 |  | 0.14 |  |
|  | **2011-2015 vs. 2016-2020** | **2011-2015** | 31/2906 (1.1%) | 3.74 (1.49-5.40) Sum=10513 | 0.29 (0.20-0.42) | 34/6964 (0.5%) | 4.81 (3.54-6.20) Sum=33043 | 0.10 (0.07-0.14) | 2.78 (1.71 - 4.53) p=<.0001 |  |  |  |
|  |  | **2016-2020** | 16/2930 (0.5%) | 3.85 (1.86-5.47) Sum=10995 | 0.15 (0.08-0.24) | 14/6805 (0.2%) | 4.82 (3.56-6.22) Sum=32542 | 0.04 (0.02-0.07) | 3.37 (1.64 - 6.91) p=0.0009 |  | 0.71 |  |
| ***Female - population*** |  |  |  |  |  |  |  |  |  |  |  |  |
| **<30 years of age at inclusion** | **2006-2010 vs. 2016-2020** | **2006-2010** | 42/696 (6.0%) | 4.94 (3.90-6.38) Sum=3511 | 1.20 (0.86-1.62) | 38/1844 (2.1%) | 5.29 (4.11-6.60) Sum=9797 | 0.39 (0.27-0.53) | 3.06 (1.97 - 4.75) p=<.0001 |  |  |  |
|  |  | **2016-2020** | 36/667 (5.4%) | 5.28 (4.04-6.70) Sum=3489 | 1.03 (0.72-1.43) | 18/1647 (1.1%) | 5.53 (4.27-6.79) Sum=9011 | 0.20 (0.12-0.32) | 5.12 (2.91 - 9.02) p=<.0001 |  | 0.16 |  |
|  | **2006-2010 vs. 2011-2015** | **2006-2010** | 42/696 (6.0%) | 4.94 (3.90-6.38) Sum=3511 | 1.20 (0.86-1.62) | 38/1844 (2.1%) | 5.29 (4.11-6.60) Sum=9797 | 0.39 (0.27-0.53) | 3.06 (1.97 - 4.75) p=<.0001 |  |  |  |
|  |  | **2011-2015** | 38/739 (5.1%) | 5.31 (3.89-6.65) Sum=3864 | 0.98 (0.70-1.35) | 44/1862 (2.4%) | 5.35 (4.07-6.75) Sum=9969 | 0.44 (0.32-0.59) | 2.22 (1.44 - 3.43) p=0.0003 |  | 0.31 |  |
|  | **2011-2015 vs. 2016-2020** | **2011-2015** | 38/739 (5.1%) | 5.31 (3.89-6.65) Sum=3864 | 0.98 (0.70-1.35) | 44/1862 (2.4%) | 5.35 (4.07-6.75) Sum=9969 | 0.44 (0.32-0.59) | 2.22 (1.44 - 3.43) p=0.0003 |  |  |  |
|  |  | **2016-2020** | 36/667 (5.4%) | 5.28 (4.04-6.70) Sum=3489 | 1.03 (0.72-1.43) | 18/1647 (1.1%) | 5.53 (4.27-6.79) Sum=9011 | 0.20 (0.12-0.32) | 5.12 (2.91 - 9.02) p=<.0001 |  | 0.021 |  |
| **30-64 years of age at inclusion** | **2006-2010 vs. 2016-2020** | **2006-2010** | 62/1570 (3.9%) | 4.71 (3.36-6.25) Sum=7131 | 0.87 (0.67-1.11) | 41/3804 (1.1%) | 5.46 (4.16-6.71) Sum=20532 | 0.20 (0.14-0.27) | 4.27 (2.88 - 6.34) p=<.0001 |  |  |  |
|  |  | **2016-2020** | 15/1158 (1.3%) | 5.08 (3.62-6.60) Sum=5665 | 0.26 (0.15-0.44) | 7/2421 (0.3%) | 5.68 (4.35-6.93) Sum=13521 | 0.05 (0.02-0.11) | 5.14 (2.09 - 12.60) p=0.0004 |  | 0.74 |  |
|  | **2006-2010 vs. 2011-2015** | **2006-2010** | 62/1570 (3.9%) | 4.71 (3.36-6.25) Sum=7131 | 0.87 (0.67-1.11) | 41/3804 (1.1%) | 5.46 (4.16-6.71) Sum=20532 | 0.20 (0.14-0.27) | 4.27 (2.88 - 6.34) p=<.0001 |  |  |  |
|  |  | **2011-2015** | 42/1395 (3.0%) | 4.96 (3.49-6.49) Sum=6673 | 0.63 (0.45-0.85) | 19/3158 (0.6%) | 5.62 (4.22-6.84) Sum=17379 | 0.11 (0.07-0.17) | 5.74 (3.34 - 9.87) p=<.0001 |  | 0.41 |  |
|  | **2011-2015 vs. 2016-2020** | **2011-2015** | 42/1395 (3.0%) | 4.96 (3.49-6.49) Sum=6673 | 0.63 (0.45-0.85) | 19/3158 (0.6%) | 5.62 (4.22-6.84) Sum=17379 | 0.11 (0.07-0.17) | 5.74 (3.34 - 9.87) p=<.0001 |  |  |  |
|  |  | **2016-2020** | 15/1158 (1.3%) | 5.08 (3.62-6.60) Sum=5665 | 0.26 (0.15-0.44) | 7/2421 (0.3%) | 5.68 (4.35-6.93) Sum=13521 | 0.05 (0.02-0.11) | 5.14 (2.09 - 12.60) p=0.0004 |  | 0.82 |  |
| **≥65 years of age at inclusion** | **2006-2010 vs. 2016-2020** | **2006-2010** | 21/1984 (1.1%) | 3.47 (1.12-5.13) Sum=6684 | 0.31 (0.19-0.48) | 23/4536 (0.5%) | 4.71 (3.49-6.11) Sum=21192 | 0.11 (0.07-0.16) | 2.83 (1.56 - 5.12) p=0.0006 |  |  |  |
|  |  | **2016-2020** | 11/1795 (0.6%) | 3.81 (1.59-5.52) Sum=6658 | 0.17 (0.08-0.30) | 10/3642 (0.3%) | 4.93 (3.64-6.40) Sum=17840 | 0.06 (0.03-0.10) | 2.88 (1.22 - 6.78) p=0.016 |  | 0.97 |  |
|  | **2006-2010 vs. 2011-2015** | **2006-2010** | 21/1984 (1.1%) | 3.47 (1.12-5.13) Sum=6684 | 0.31 (0.19-0.48) | 23/4536 (0.5%) | 4.71 (3.49-6.11) Sum=21192 | 0.11 (0.07-0.16) | 2.83 (1.56 - 5.12) p=0.0006 |  |  |  |
|  |  | **2011-2015** | 19/2017 (0.9%) | 3.67 (1.39-5.28) Sum=7122 | 0.27 (0.16-0.42) | 24/4281 (0.6%) | 4.88 (3.62-6.32) Sum=20654 | 0.12 (0.07-0.17) | 2.29 (1.25 - 4.19) p=0.0071 |  | 0.59 |  |
|  | **2011-2015 vs. 2016-2020** | **2011-2015** | 19/2017 (0.9%) | 3.67 (1.39-5.28) Sum=7122 | 0.27 (0.16-0.42) | 24/4281 (0.6%) | 4.88 (3.62-6.32) Sum=20654 | 0.12 (0.07-0.17) | 2.29 (1.25 - 4.19) p=0.0071 |  |  |  |
|  |  | **2016-2020** | 11/1795 (0.6%) | 3.81 (1.59-5.52) Sum=6658 | 0.17 (0.08-0.30) | 10/3642 (0.3%) | 4.93 (3.64-6.40) Sum=17840 | 0.06 (0.03-0.10) | 2.88 (1.22 - 6.78) p=0.016 |  | 0.64 |  |
| Confidence interval for unadjusted event rates per 100 person years are obtained from exact Poisson confidence limits. Cox regression was used for time to any event presented by HR. * Adjusted for Age at inclusion, Sex, Stroke, TBI, Dementia, Brain tumour, Brain infection, Diabetes, Cardiovascular diseases, Intellectual disability, Cancer, Specialist psychiatric care (before date of inclusion). The p-value for interaction was calculated between study group and each pairwise time period. | | | | | | | | | | | | |

### **eTable 5. Risk factors for new-onset depression from Cox regression during periods 2006-2010, 2011-2015 and 2016-2020 (FAS population - Epilepsy cases**

|  | | **Patients with epilepsy N=28569** | | | | | |  |
| --- | --- | --- | --- | --- | --- | --- | --- | --- |
|  | | **2006-2010 N=9561** | | **2011-2015 N=9843** | | **2016-2020 N=9165** | |  |
| **Variable** | **Comparison** | **HR (95% CI)** | **p-value** | **HR (95% CI)** | **p-value** | **HR (95% CI)** | **p-value** | **p-value interaction** |
| **Sex** | **Male vs Female** | 0.83 (0.76 - 0.91) | <.0001 | 0.82 (0.75 - 0.90) | <.0001 | 0.81 (0.74 - 0.89) | <.0001 | 0.73 |
| **Age at inclusion** | **Per 1 year increase** | 1.01 (1.01 - 1.01) | <.0001 | 1.01 (1.01 - 1.01) | <.0001 | 1.01 (1.01 - 1.01) | <.0001 | 0.74 |
| **Stroke** | **Yes vs No** | 1.94 (1.77 - 2.12) | <.0001 | 1.64 (1.49 - 1.80) | <.0001 | 1.65 (1.50 - 1.81) | <.0001 | 0.012 |
| **TBI** | **Yes vs No** | 1.20 (1.01 - 1.44) | 0.040 | 1.27 (1.08 - 1.50) | 0.0045 | 1.33 (1.12 - 1.59) | 0.0011 | 0.43 |
| **Dementia** | **Yes vs No** | 1.38 (1.11 - 1.70) | 0.0032 | 1.62 (1.35 - 1.96) | <.0001 | 1.80 (1.48 - 2.20) | <.0001 | 0.09 |
| **Brain tumour** | **Yes vs No** | 1.32 (1.14 - 1.52) | 0.0001 | 1.30 (1.13 - 1.49) | 0.0002 | 1.58 (1.38 - 1.80) | <.0001 | 0.09 |
| **Brain infection** | **Yes vs No** | 1.14 (0.81 - 1.61) | 0.45 | 1.54 (1.19 - 1.99) | 0.0012 | 1.31 (0.97 - 1.77) | 0.07 | 0.59 |
| **Diabetes** | **Yes vs No** | 1.30 (1.13 - 1.51) | 0.0004 | 1.46 (1.29 - 1.65) | <.0001 | 1.31 (1.15 - 1.50) | <.0001 | 0.95 |
| **Cardiovascular diseases** | **Yes vs No** | 1.55 (1.42 - 1.70) | <.0001 | 1.60 (1.47 - 1.75) | <.0001 | 1.51 (1.38 - 1.65) | <.0001 | 0.64 |
| **Intellectual disability** | **Yes vs No** | 0.60 (0.41 - 0.86) | 0.0061 | 0.80 (0.59 - 1.08) | 0.14 | 0.57 (0.41 - 0.79) | 0.0009 | 0.78 |
| **Cancer** | **Yes vs No** | 1.37 (1.20 - 1.56) | <.0001 | 1.47 (1.30 - 1.66) | <.0001 | 1.66 (1.47 - 1.87) | <.0001 | 0.053 |
| Cox regression was used for time to the event of new-onset depression, defined as ATC code N06A, presented by HR. The p-value for interaction was calculated for each covariate, as the interaction between the covariate across the time periods. | | | | | | | | |
